# Quantum-enhanced nanodiamond rapid test advances early SARS-CoV-2 antigen detection in clinical diagnostics

**DOI:** 10.1101/2025.06.19.25329920

**Authors:** Alyssa Thomas DeCruz, Benjamin S. Miller, Da Huang, Max McRobbie, Felix Donaldson, Laura E McCoy, Ciara K. O’Sullivan, Johannes C. Botha, Eleni Nastouli, Rachel A. McKendry

## Abstract

Worldwide, the urgent need for more sensitive rapid lateral flow tests (LFTs) for early disease diagnosis is driving advances in quantum technologies. Spin-enhanced nanodiamond LFTs offer the potential for a sensitivity step change, yet to date only model systems have been studied with few clinical samples. Herein, we report the largest spin-enhanced LFT clinical evaluation, focusing on SARS-CoV-2 antigen as an exemplar. The limit of detection for recombinant antigens was 0.67 pg/ml, and inactivated whole virus wild-type and Omicron variants were 13 TCID_50_/mL and 8.8 TCID_50_/mL, respectively. Our blinded clinical study with 103 patient upper respiratory tract swab samples showed 95.1% sensitivity (Ct ≤ 30) and 100% specificity benchmarked to RT-qPCR, with no cross-reactivity to influenza A, RSV, and Rhinovirus. Analysis of trial data indicates spin-enhanced LFTs could diagnose SARS-CoV-2 on average 2.0 days earlier than conventional gold nanoparticle LFTs with identical antibodies, and just 0.6 days after RT-qPCR. Quantum-enhanced LFT sensitivity would have detected ∼69,400 more patients in a single day at the peak of Omicron, reducing the transmission risk and protecting populations. Our findings mark an important milestone in the emerging field of quantum-enhanced diagnostics, with the potential for significant health-economic benefits to patients, populations, health systems and society.

## Main

The field of quantum technologies for biomedical applications has experienced remarkable growth in recent years. Quantum sensors, in particular, have emerged as a transformative technology, offering enhancements in sensitivity, spatial and temporal resolution enabled by the interaction of biochemical targets with single or ensemble quantum systems^1^. One of the primary systems being studied for biomedical sensing is the nitrogen-vacancy centre (NV) in diamond^2^. This optically active, defect-bound spin system maintains coherence at room temperature and exhibits spin dependent fluorescence emission, enabling optical readout of the spin state as well as controllable modulation of fluorescence. The NV Hamiltonian is sensitive to a broad range of physical parameters, while the defects can be hosted in bulk diamond or diamond nanoparticles, leading to a broad set of biomedical sensing applications including; nanoscale nuclear magnetic resonance spectroscopy^3^, dynamic particle orientation tracking^4^ as well as pH and temperature sensing in cells^5^. Zeeman splitting of the sublevels by external magnetic fields has been utilised in the imaging of cells^6^ and individual analytes^7,8^. Fluorescent nanodiamonds (FND) containing NV centres have also been studied for *in vivo* applications, enabling nanoscale tracking of metabolic processes within living cells^9^.

FNDs have also found exciting applications in in-vitro diagnostics such as lateral flow tests (LFTs). Fluorescent labels are commonly used to improve test sensitivity, but often suffer from bleaching, blinking, and are limited by variable background autofluorescence^10,11^. FNDs hold potential to overcome these limitations, owning to their high brightness, photostability, and the ability to selectively modulate the signal^12,13^. This is particularly useful in LFTs where the performance of fluorescent reporter particles is often limited by background autofluorescence from the nitrocellulose membrane^14^. Our recent work demonstrated the potential of spin-enhanced FNDs in a rapid LFT, achieving a fundamental detection limit of 8.2 x 10^-19^ M, 94,000-fold more sensitive than conventional gold nanoparticles^15^. In a diagnostic assay, however, this limit is not reached because of nonspecific binding of nanoparticles to the membrane, reflected by the 7,500-fold sensitivity improvement achieved for the HIV-1 RNA model assay in the same work^15^.

Although the FND platform is broadly target agnostic, Miller et al. ^15^ largely focused on nucleic acid detection, achieving single-copy amplicon detection using an isothermal amplification step. In direct antigen detection assays, where target amplification is not possible, nanoparticle detection limits translate directly to analytical sensitivities. Consequently, spin-enhanced FND biosensors could be impactful in addressing the need for more sensitive antigen tests for use in resource limited settings such as doctors’ surgeries, pharmacies and even self-testing at home. Key challenges for FND-based antigen tests are sensitivity and clinical evaluation. For example, Wei-Wen Hsiao et al.’s^16^ limit of detection (LoD) of 1.94 ng/mL for SARS-CoV-2 nucleocapsid protein remains comparable to simpler gold nanoparticle-based LFTs. In later work, Le et al.^17^, achieved a LoD of 0.02 ng/mL detecting ESTAT6 for tuberculosis diagnosis but direct antigen detection was limited by the use of cell-cultured samples. In a non-modulated FND approach, Feuerstein et al.^18^ demonstrated the feasibility of FND-based LFTs for Ebola glycoprotein detection but encountered sensitivity limitations in serum samples. Notably, sensitivity remains a key challenge and no FND studies have been reported using direct (non-cultured) clinical samples, with the exception of our proof-of-concept study, Miller et al.^15^, where we tested a limited number of clinical samples: one clinical standard plasma sample and a seroconversion panel for the HIV-1 RNA assay. Hence, there remain significant research challenges translating sensitivity in model systems through to clinical samples^18^, highlighting the need for large-scale clinical evaluation and clinical benchmarking to gold standard diagnostic methods such as PCR.

The development of antigen LFTs is a key component of the WHO R&D Blueprint strategy to address priority diseases with epidemic potential^19^ such as coronaviruses, Crimean-Congo haemorrhagic fever and Ebola where sensitivity is a challenge. Additionally, the Foundation for Innovative New Diagnostics has just published a new Pathogen Diagnostic Readiness Index indicating large unmet needs in diagnostic performance across diseases of epidemic potential (www.finddx.org/data-and-impact/dashboards/diagnostic-readiness-index). Herein, we aim to develop an FND antigen assay to deliver a step change in sensitivity to early-stage infections, and undertake a large, blinded clinical evaluation, focusing on SARS-CoV-2 as an exemplar. In 2019, SARS-CoV-2, a previously unknown virus, emerged infecting over 700 million people globally, leading to >7 million deaths by 2024 (www.data.who.int/dashboards/covid19/cases). SARS-CoV-2 is characterised by its high transmissibility, long incubation period, and high number of asymptomatic carriers^20^ - key factors that led to accelerated disease spread and subsequent pandemic. Consequently, timely diagnostics played a key role in the identification of cases to reduce community transmission^21^. Nucleic acid amplification tests (e.g. PCR tests) targeting viral RNA were quickly adopted as the gold standard for COVID-19 diagnosis^22^, with antigen-detecting LFTs (Ag-LFTs) later implemented for widespread use at the point-of-care or home settings^23^. Confirmatory testing for COVID-19 has relied on quantitative reverse transcriptase PCR (RT-qPCR) due to its high sensitivity and specificity towards SARS-CoV-2 genes in respiratory tract specimens^24^. The detection limits of RT-qPCR can reach as low as 1-100 copies/mL, useful for early detection in the first 24-48hrs of infection (Fig. 1a)^25^. Despite the high sensitivity, molecular methods are inherently difficult to scale-up as seen during the COVID-19 pandemic, when RT-qPCR was unable to meet high frequency testing demands due to high cost, personnel, sample transportation and long turn-around times of 1-2hrs^26^. Ag-LFTs were widely adopted as an alternative to molecular methods to meet the high testing demand^27^.

**Fig. 1:**
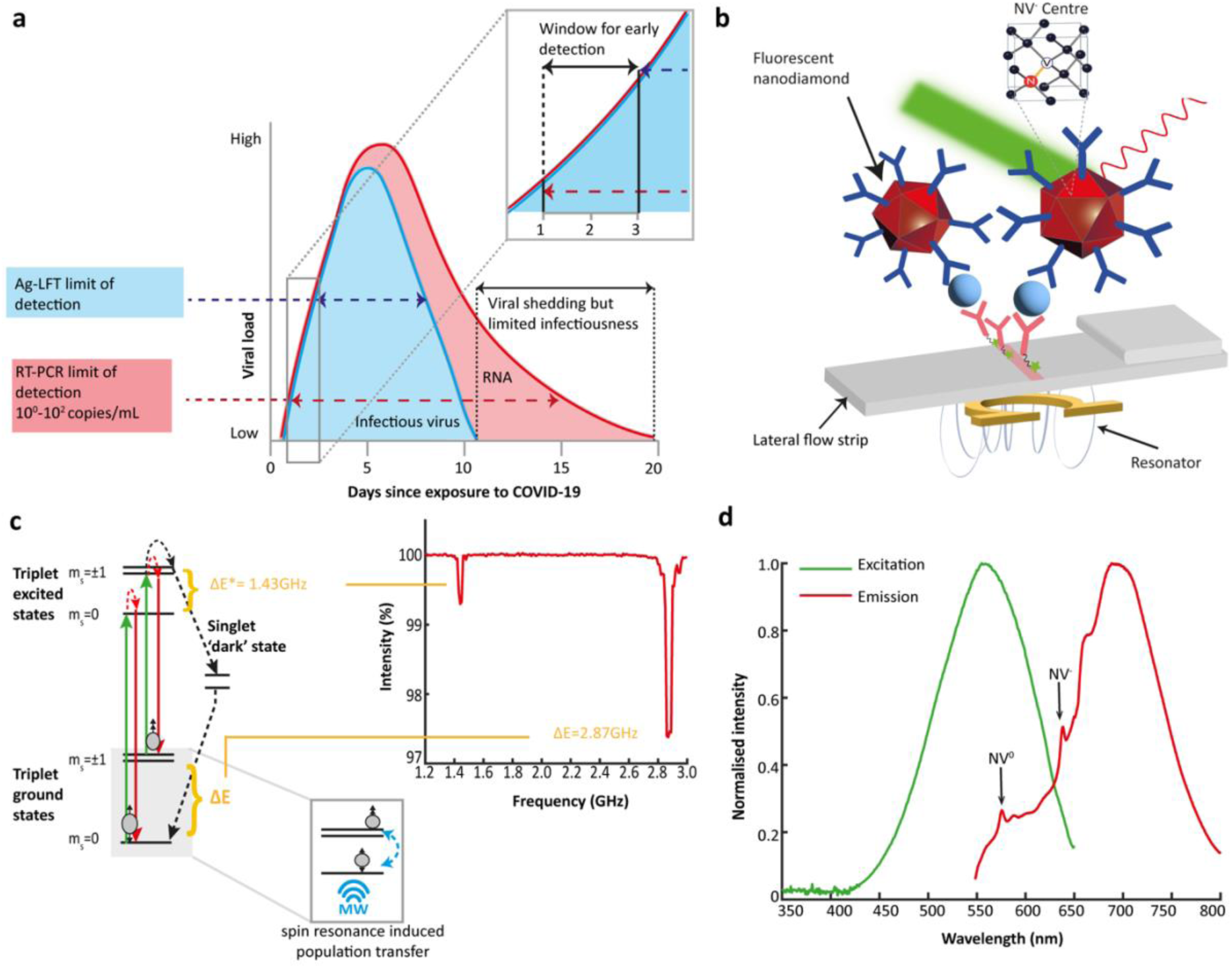
Early detection of SARS-CoV-2 antigen by spin-enhanced LFT and FND characterisation. **(a)** SARS-CoV-2 infection dynamics of RNA and infectious virus by viral load with the LoD of RT-qPCR at ∼10^0^-10^2^ copies/mL, detecting active virus 1-3 days earlier than current antigen-detecting LFTs. Call-out box shows the 1-3 days window for early detection with improved sensitivity of Ag-LFTs. **(b)** Schematic of FNDs immobilised at the LFT test line via antibody sandwich complex. The FND is excited at 550nm and emits from the visible to the NIR. A small omega-shaped resonator is driven at the NV^-^ zero-field splitting frequency (2.87 GHz) to induce spin population transfer between the m_s_=0 and m_s_=±1 states. The amplitude of this driving field is modulated to provide a time varying fluorescence signal. **(c)** Energy level diagram of NV-centre, showing the zero-field splitting of the spin triplet optical ground and excited states, the spin conserving optical transitions, and the non-radiative decay pathway from the optically excited m_s_=±1 state via the metastable singlet states. Under continuous optical illumination, this non-spin-conserving decay pathway results in an increase in population in the m_s_=0 state in a process known as optical initialisation. Once a population difference between the m_s_=0 or m_s_=±1 is established, if energy resonant with the spin transitions is provided (ΔE*= 1.43GHz and ΔE= 2.87GHz), population is transferred from the m_s_=0 to the m_s_=±1 states. Due to the singlet decay pathway not resulting in a visible photon, this reduces the detected photoluminescence. This effect is demonstrated by the results of the continuous wave optically detected magnetic resonance (CW-ODMR) experiment shown in the inset with reductions in fluorescence centred at ΔE*and ΔE. **(d)** Excitation and emission spectra showing an excitation peak at 550nm and emission peak at 675nm

Ag-LFTs target the spike or nucleocapsid viral protein detected from nasal or throat swab sample without the need for additional extraction steps^28^. SARS-CoV-2 Ag-LFTs are low cost, fast (<15mins), and amenable non-traditional healthcare settings^23^. Importantly, antigen positivity correlates well with infectiousness and may therefore hold advantages over RT-qPCR which faces prolonged RNA positivity^29^ (Fig. 1a). Yet despite the development of over 1,000 commercial antigen tests (www.finddx.org/covid-19/test-directory/), the majority still lack sensitivity, with limits of detection equating to ∼10^5^-10^6^ copies/mL or Cts<25^30,31^. Furthermore, many LFTs see a significant decline in performance in moderate viral load samples (Ct 25 to 30)^32^ and hence positive cases could be missed, with detection occurring up to 1-2 days later than RT-qPCR (Fig. 1a). This window for early detection relies on both high sensitivity and rapid turn-around time, which quantum sensing holds potential to address.

## Results and discussion

These results describe the development of a SARS-CoV-2 antigen FND Ag-LFT assay and its evaluation by a large, blinded clinical evaluation.

### Spin-enhanced fluorescent nanodiamond characterisation

Antibody-functionalised FNDs are used as the detection nanoparticle forming a sandwich complex between the target analyte and secondary capture antibody (Fig. 1b). Size and morphology of 600nm FND-PG were observed by SEM (Extended Data Fig. 1a).

Fig. 1c shows the energy level diagram of an NV^-^ centre with spin triplet ground and optically excited states, and a pair of intermediate metastable singlet dark states. Green light (550nm) excites electrons into the optically excited manifold, which can then decay back down to the ground state, emitting a photon (∼675nm), where spin is conserved throughout. Alternatively, electrons in the m_s_=±1 excited state can decay via the singlet ‘dark’ state, where no visible photon is emitted, in a non-spin-conserving relaxation process. This enables the spin sub-levels to be distinguished via fluorescence intensity - optically detected magnetic resonance (ODMR) - as well as optical initialisation of the population into the m_s_ = 0 state. Once initialised, controllable reduction of the photoluminescence can then be achieved by transferring population from the m_s_=0 to the m_s_=±1 states (increasing the spin population decaying via the dark state) through the application of a driving field with energy resonant with either the ground (ΔE= 2.87GHz) or optically excited (ΔE*= 1.43GHz) zero-field splitting. This reduction in fluorescence is evident in the ODMR spectrum of FNDs immobilised at the test line of a nitrocellulose lateral flow strip in Fig. 1c. Fluorescence is reduced by 0.70% at 1.43GHz and 2.6% at 2.87GHz, corresponding to the excited (ΔE*) and ground state (ΔE) zero field splitting, respectively. Selectively controlling fluorescent intensity provides the ideal mechanism for lock-in detection as the effect is highly specific to the fluorescent source. Fluorescent intensity scales with number of NV centres per particle^15^, therefore larger particles ∼600nm FNDs were selected for this work. In addition, the optical spectrum of NV^-^ centres (Fig. 1d) is well-suited for biosensing applications, away from most biological autofluorescence, which is at shorter wavelengths^33^.

### Design of FND-based LFT for SARS-CoV-2 antigen detection

Sourcing high affinity capture reagents is critical to diagnostic sensitivity and specificity, but remains a challenge due to high cost^34,35^. Receptor-ligand binding kinetics and mass transport impact LFT performance, where the ratio of signal from specific analyte-mediated binding to non-specific binding and background signal should be optimised^36^. Poor kinetics limit sensitivity by reducing the fraction of bound antigen, reducing signals. While increasing nanoparticle concentration can increase specific binding rates, it also increases non-specific binding^15^.

We identified six antibodies: four commercially available SARS-CoV-2 nucleocapsid antibodies (40143-R001, 40143-R004, 40143-R040, 40143-MM08; Sino Biological) with reported high affinities (K_D_≍0.02nM), plus two in-house SARS-CoV antibodies (CR3018 and CR3009; UCL, Division of Infection and Immunity). We used Biolayer-interferometry to measure antibody binding affinity (K_D_) (Fig. 2a) and calculate rate of association (k_on_) and dissociation (k_off_). We found the K_D_ values ranged from 0.19 nM to 69 nM and k_on_ values 1.2 - 3.6×10^4^ s^-1^nM^-1^. Antibody MM08 (AbMM08) had the highest K_D_ of 0.19 nM [95% CI: 0.091-0.36] and the fastest k_on_ of 3.6×10^5^ s^-1^M^-1^ [95% CI: 3.2-3.9×10^5^]. We proceeded with all four commercial antibodies, found to have stronger binding affinities (K_D_ values 0.19 nM to 0.34 nM), and for their demonstration as high performance antibodies in literature. This included AbMM08, AbR004, and AbR001 ranking amongst the top 5 performing antibody pairs in a large evaluation of 1021 SARS-CoV-2 antibody pairs (Cate et al.^37^) and numerous early reports establishing their high quality during the pandemics initial test development phases^38–40^. Note, antibodies CR3018 and CR3009 were provided early on, previously developed against SARS-CoV^41^, whereas the commercial antibodies were later developed specifically against SARS-CoV-2 hence higher binding affinity can also be attributed to specificity to the target antigen. In our data, antibodies R001 and R040 showed significantly weaker binding than manufacturer reports. Kinetic parameters are detailed in Supplementary Information Fig.1 and Supplementay Information Table 1.

**Fig. 2:**
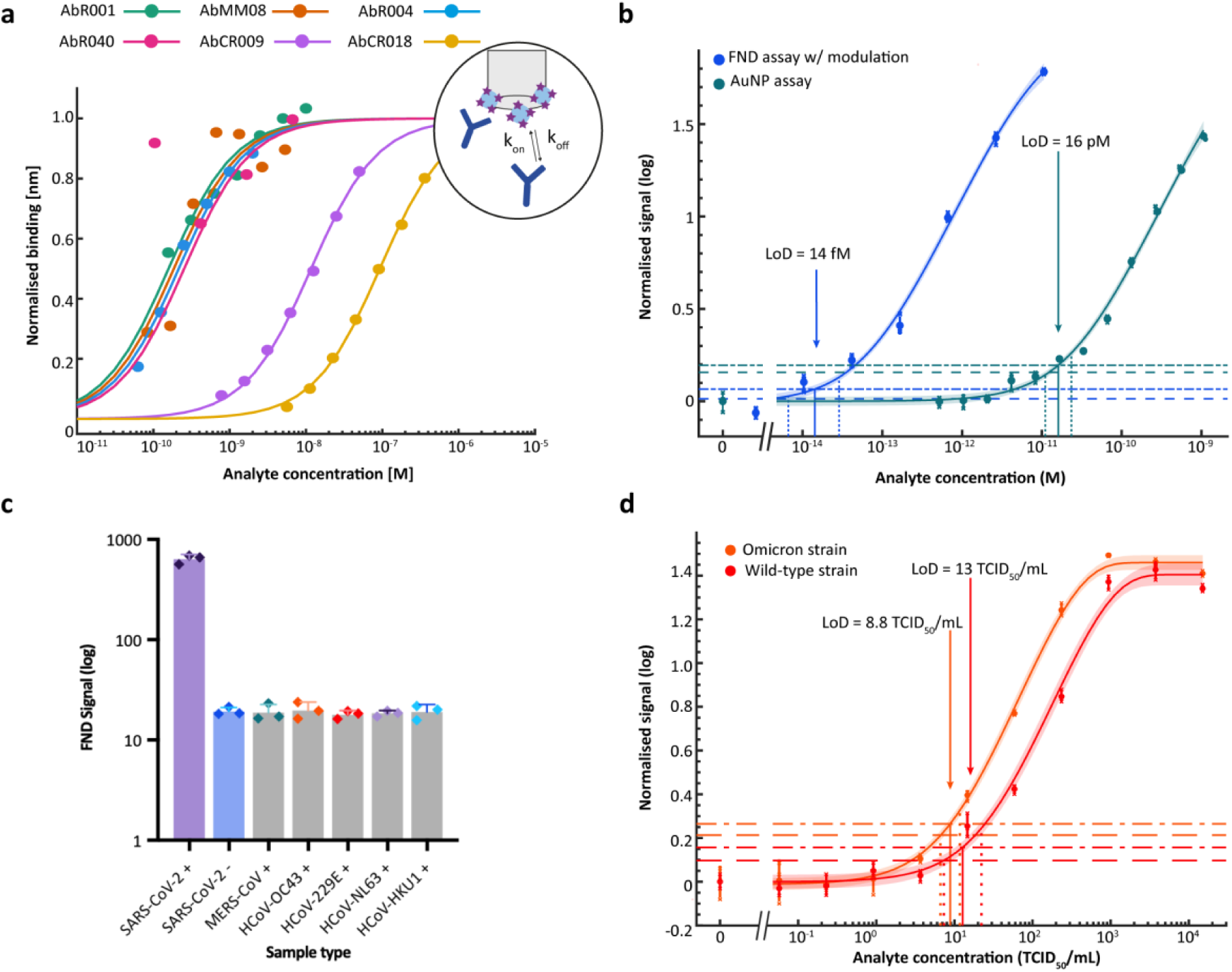
Antibody characterisation and FND Ag-LFT assay analytical sensitivity and specificity. **(a)** Fitted curves of SARS-CoV-2 antibody-antigen binding over a range of antigen concentrations evaluated using Biolayer-interferometry. Sensor design shown in schematic. **(b)** Serial dilution of recombinant nucleocapsid protein with FND Ag-LFT evaluated by lock-in modulation analysis (blue) resulting in a LoD of 14 fM [95%CI: 6.6-29] using a Langmuir fitting method. This was compared to 40nm gold nanoparticles (green) with an LoD of 16 pM [95%CI: 11-23]. Dots show mean and error bars of n=3 replicates. **(c)** Mean FND signal (n=3) of recombinant antigens from other human coronaviruses, showing no statistically significant difference from SARS-CoV-2 negatives (p-value>0.9999, one-way ANOVA, F=0.19, n=18, post-hoc Dunnett’s) **(d)** Serial dilution of SARS-CoV-2 gamma-irradiated whole virus of both Wild-type (red) and Omicron (orange) strains evaluated by lock-in analysis showing an LoD of 13 TCID_50_/mL [95% CI: 7.3-22] and 8.8 TCID_50_/mL [95% CI: 6.6-12], respectively.

A step-wise approach was used to screen antibody pairs based on optimised binding interactions where first, the binding affinity of the detection antibody was measured, followed by the secondary antibody binding affinity in a sandwich format, minimizing epitope binding interference and non-specific binding. The antibody-functionalised FNDs were first evaluated in direct bind format, targeting biotinylated recombinant nucleocapsid protein. The strong binding affinity of biotin-avidin ensures effective capture at the test line, making the FND-antibody complexation with the analyte the limiting interaction. The signal-to-noise ratio (SNR) is defined as the ratio of the lock-in amplitude of a positive test line compared to the mean lock-in amplitude of the negative controls, where the signal is due to FNDs non-specifically bound to the membrane, and variation is caused by strip-to-strip variation. Here the SNR of a strong positive sample (10ng/mL) was used to evaluate antibody-conjugate performance. FND-AbMM08 showed the highest SNR of 84 although there were no significant differences (p-value=0.7058, ANOVA, F=0.48, df=11, post-hoc Tukey’s multiple comparisons), shown in Extended Data Fig. 1b. FND-AbMM08 was selected as the best performing detection antibody for the FND system.

Functionalisation and dispersity of the FND conjugate was measured using dynamic light scattering. The 600nm FND-PG showed average hydrodynamic diameter at 628nm (±220) and a polydispersity index (PDI) of 24%, consistent with approximate size of particles with a polyglycerol coating (Extended Data Fig. 1c). FNDs functionalised with AbMM08 showed a 20nm increase in hydrodynamic diameter to 643nm (±140) and PDI of 21%, corresponding to the approximate theoretical size of a typical antibody (15nm). Secondary capture antibody pairs tested with FND-AbMM08 showed that AbR001 out-performed AbR004 (p-value=0.025) and AbR040 (p-value=0.036, ANOVA, F=8.2, df=8, post-hoc Tukey’s multiple comparisons) (Extended Data Fig. 1d). Therefore, FND-AbMM08 and capture AbR001 was used for further assay optimisation and performance evaluation. It is important to note that the antibody pair evaluation was conducted using a consistent recombinant antigen and potential antibody pair selection bias resulting from screening on a single antigen source was not considered in this initial down-selection process^37^. Antibody-printed test lines require high volumes and a large amount of excess antibody therefore we used biotinylated AbR001 (AbR001-b) in solution with polystreptavidin-printed test line to reduce antibody consumption by 84.6% and therefore cost.

The concentration of FND-Ab and capture antibody, and buffer formulations were then optimised. To ensure optimal binding, a high concentration of capture antibody AbR001-b was used to screen through FND-AbMM08 conjugate concentrations ranging from 6.6 fM to 42 pM. The positive and negative signal scaled linearly with increasing FND concentration where the SNR, found by dividing the fitted linear regressions, resulted in a constant value of ∼20. A concentration of 26 fM was selected to minimise reagent consumption while also generating a measurable intensity for the strong positive test line (Extended Data Fig. 2a,b). Buffer components were re-formulated to suit clinical testing of SARS-CoV-2 nasal swabs, which requires viral lysis components in addition to non-specific protein blockers. Final buffer composition found the addition of 0.8% casein and 0.05% Tween20 improved the SNR and IGEPAL CA-630 was found to be an effective detergent for viral lysis (Extended Data Fig. 2c,d).

### Analytical sensitivity of spin-enhanced FND LFT compared with gold nanoparticles

The sensitivity of the FND LFT was first evaluated using a serial dilution of SARS-CoV-2 recombinant nucleocapsid protein where the lock-in fluorescent intensity was used to plot and quantify the analytical LoD using previously reported statistical method applying an exponential curve fit^42^. Comparison of different fitting models are shown in Supplementary Information Table 2. The assay here presented yielded an LoD of 14 fM [95% CI: 6.6-29] (0.67 pg/mL) with spin-modulated lock-in (Fig. 2b). Note that the signal is also limited by non-specific binding of FNDs to the nitrocellulose - FND signal can still be detected in the negative sample (Extended Data Fig. 3a). To fully exploit the detection sensitivity of spin-enhanced FND detection, non-specific binding should be further reduced below the readout noise floor.

The sensitivity of the FND assay was then benchmarked to that of a conventional AuNP-based assay developed in-house using the same reagents. The AuNP assay yielded an LoD of 16 pM [95% CI: 11-23], demonstrating a 1,100-fold sensitivity improvement with FNDs (Fig. 2b, p-value<0.0001, t-test, T=-18, df=70). Test line images are shown in Extended Data Fig. 3b. The analytical sensitivity reached in this work demonstrates superior sensitivity of spin-enhanced LFTs compared to similar low-cost rapid LTFs reported in literature^16,38,43–45^ (Supplementary Information Table 3) and reaches LoDs comparable to high-throughput, microarray platforms such as Quanterix’s Simoa. This platform reports a lower limit of quantification (LLOQ) of 0.31 pg/mL and LoD of 0.099 pg/mL [0.046-0.20] but at a substantially higher cost of ∼£400,000 for the HDx platform and £2,400 per SARS-CoV-2 N protein kit for up to 96 samples. This is compared to an approximate cost of £1.30 per FND strip and a low-cost reusable reader without requiring specialised technologists^15^.

The specificity of the FND assay to SARS-CoV-2 was tested against recombinant antigens of other common coronaviruses. No cross-reactivity was observed for MERS-CoV (p-value=0.99), HCoV-OC43 (p-value=0.99), HCoV-299E (p-value=0.95), HCoV-NL63 (p-value=0.99) and HCoV-HKU1 (Fig. 2c, p-value>0.9999, one-way ANOVA, F=0.19, df=17, post-hoc Dunnett’s relative to SARS-CoV-2 negatives). Next, the technical performance was further demonstrated in a more clinically relevant sample, using a serial dilution of the whole inactivated virus samples in VTM. The assay showed the ability to detect both the Wild-Type and Omicron variants with similar sensitivities, yielding LoD’s of 13 TCID_50_/mL [95% CI: 7.3-22] and 8.8 TCID_50_/mL [95% CI: 6.6-12], respectively (Fig. 2d, p-value=0.26, t-test, T=1.2, df=45). This affirms the choice of targeting the highly conserved nucleocapsid protein whereby performance is less likely to be compromised by emerging variants ^46^. Furthermore, the viral RNA concentration (ddPCR) of the stock viral isolate enabled the conversion of estimated LoD correlating to ∼3 x 10^4^ genome copies/mL, which indicates a better LoD than most LFTs on the market with LoDs as high as 10^5^-10^6^ genome copies/mL ^47^. For comparison to a widely used commercially available rapid LFT, FlowFlex assay showed a visible test line around 4 x 10^3^ TCID_50_/mL using the wild-type viral isolate tested in-house, which is ∼13-fold lower than the claimed sensitivity of ∼300 TCID_50_/mL and 300-fold less sensitive than the FND assay above (Extended Data Fig. 3c). However, this comparison is less pertinent than the comparison with the in-house gold nanoparticle assay (Fig. 2b), as commercial tests likely use different antibodies, buffers, and membranes, all of which will affect sensitivity, confounding the nanoparticle comparison.

### Blinded study to determine clinical sensitivity of FND Ag-LFT compared to RT-qPCR

An important next step was to blindly evaluate the performance of the FND assay using clinical swab samples compared to gold standard RT-qPCR results (Ct value). 103 frozen samples, a combination of nasopharyngeal and nasal swabs, were received from UCLH collected from individuals (returning travellers and symptomatic patients) from Nov-Dec 2022 and Jun-Aug 2023 during the pandemic. The initial sample set was selected at random by UCLH to broadly represent patient sample pool viral loads. A further set of samples was requested to analyse the lower limits of detection i.e. high Ct values. The samples comprised 53 SARS-CoV-2 positive samples with Ct values between 17 and 37, and 50 SARS-CoV-2 negative samples, 13 of which were positive for other respiratory viruses including Flu A, RSV, and RHINO virus. An in-house RT-qPCR standard curve was used to convert Ct values to RNA copies/mL. Sensitivity was determined using ROC analysis with 37 SARS-CoV-2 negative swab samples as the negative control to determine the threshold cut-off. Sensitivity was analysed by grouped Ct values commonly used in literature (Fig. 3a). The FND assay showed positive correlation (correlation coefficient of 0.87 [95% Bayesian credible interval: 0.81, 0.91]) with RNA concentration, detecting samples with concentrations ≥10^4^ copies/mL (Fig. 3b). The assay maintained 100% sensitivity in high viral load samples (Ct≤25) and 86.8% sensitivity (area under curve: 0.92) across the whole Ct range (17-37) (Fig. 3c, Supplementary Information Section 4a). No significant differences in FND signals or viral loads were found between nasopharyngeal and nasal swabs (t-test p-values 0.6457 and 0.9886, respectively) as shown in Supplementary Information Fig. 3). The assay showed high specificity of 100% (n=50) and found no cross-reactivity in swab samples confirmed SARS-CoV-2 negative and positive for other common respiratory viruses including Flu A (n=6), RSV (n=6) and RHINO virus (n=1). One-way ANOVA with post-hoc Dunnett’s determined no significant differences from mean SARS-CoV-2 negative controls (all p-values>0.9999), and significant differences from the mean SARS-CoV-2 positive (p-value<0.0001, F=9.4, df=103) (Fig. 3d). Only one RHINO virus sample was included in this evaluation so statistical significance could not be determined for this category.

**Fig. 3:**
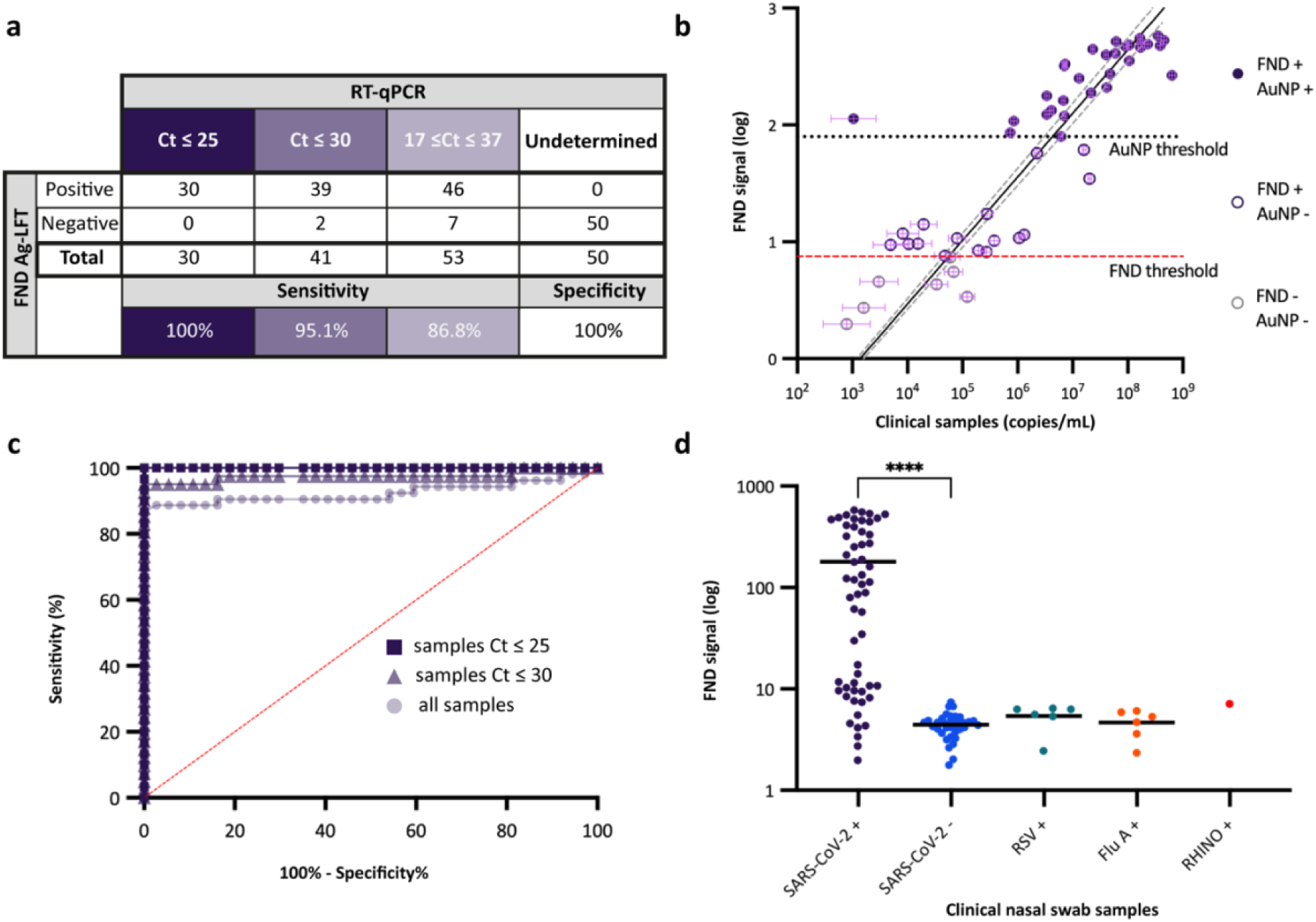
Clinical evaluation of FND Ag-LFT assay. **(a)** Sensitivity and specificity evaluation of FND Ag-LFT compared to RT-qPCR grouped by sample Ct values. Sensitivity= 100%, 95.1%, and 86.8% for Ct≤25, Ct≤30, and all samples, respectively. Specificity was 100% **(b)** Raw data plot of 53 SARS-CoV-2 positive clinical nasal swab samples with FND-Ag test line signal plotted against clinical sample concentrations (copies/mL) determined by RT-qPCR (Extended Data Fig.4). The data was fitted to a symmetrical Bayesian linear regression with errors-in-variables in x and y^58^ to account for uncertainty in copies/mL quantification from RT-qPCR and FND signal (Extended data Fig.5; Supplementary Information Section 6). Purple indicates FND true positive, purple outline indicate FND positive and AuNP negative, and grey outline indicates RT-qPCR positive and negative on the FND-Ag assay and AuNP assay. Red dashed line indicates FND Ag-LFT cut-off threshold set to give false positive and false negative rates of 5% (n=37). The black dotted line indicates extrapolated threshold for AuNP assay (Supplementary Information Section 5) **(c)** ROC analysis of clinical swab samples using 37 SARS-CoV-2 negative control samples and 53 positive samples showing sensitivity and specificity trade-off across all samples and sub-categories of samples with Ct ≤ 25 and Ct ≤ 30. See full ROC parameters in Supplementary Information Section 4a **(d)** Raw data plot of clinical nasal swab samples positive for other respiratory viruses. Bar represents mean signal. One-way ANOVA and Dunnett’s post-hoc test used to determine significance from SARS-CoV-2 negative samples, **** denotes p-value <0.0001 for SARS-CoV-2 positives, p-value>0.9999 for RSV+, FluA+, and RHINO virus+.

Importantly, this sensitivity exceeds the WHO target product profile criteria for SARS-CoV-2 RDTs with “acceptable” sensitivity of ≥80% for Ct ≍ 25 or 10^6^ genome copies/mL and “desirable” sensitivity ≥90% for Ct ≍ 30 or 10^4^ genome copies/mL, with 100% sensitivity in Cts≤25 and 95.1% in Cts≤30 (Fig. 3c). Many commercially available Ag-RDTs, including FlowFlex SARS-CoV-2 Ag (ACON Biotech) and BD Veritor system for rapid detection of SARS-CoV-2 (Becton Dickinson), lack sensitivity in samples of Ct>25, where an evaluation of 122 CE-marked SARS-CoV-2 Ag RDTs found only 20.8% (20 tests) showed a detection rate >75% in samples of Ct range 25-30 ^47^. Commercial LFTs are proprietary and have many differing parameters that can impact test performance, one of these being the antibody pairs. Therefore, we also evaluated the expected performance of our in-house AuNP assay with the same antibody pairs to serve as a direct comparator against FND performance.

For comparison, the clinical sensitivity of the AuNP assay was calculated using the fitted exponential model of the recombinant antigen data (Fig 2b and Supplementary Information Section 5) to extrapolate the AuNP limit of detection in terms of FND signal. This yielded a fitted AuNP clinical sensitivity of 56.6%, where only 2/10 moderate viral load (∼10^4^-10^5^ copies/mL) samples may be detected (Fig. 3b, Supplementary Information Table 5). The fitted AuNP assay threshold of ∼10^6^ copies/mL aligns well with sensitivity of commercially available LFTs reported in literature^30–32^ and independent evaluations such as FINDDx (https://www.finddx.org/covid-19/). These relative thresholds, and clinical sample viral load concentrations were taken together with infection dynamics reported in literature^25,48,49^ to calculate the FND diagnostic advantage in terms of days before diagnosis (Supplementary Information Section 6). Using human challenge data^25^, the patients could be diagnosed 2.0 days earlier, on average [95% credible interval: 1.8 to 2.1, P(days_FND_>days_AuNP_)<10^-10^ (25 standard errors from zero)] using the FND assay, compared to AuNPs (Fig. 4a). The FND assay could detect 95% of patients with 95% confidence no later than 3.5 days, compared to 6.2 days for the AuNP assay, a diagnostic advantage of 2.8 days (Fig 4a,b). The human challenge SARS-CoV-2 infection data-where a person in inoculated with the virus-is the only study format where the true day of initial infection is known, independent of symptom onset, but is limited by the sample size of n=18. Our sensitivity analysis was then extended to a larger clinical study (Frediani et al.^48^) that provides patient data (n=338) in grouped Ct ranges per day following symptom onset. This suggests that ∼117% (2.2-fold) more patients could be diagnosed with the FND assay on the first day of symptom onset than the conventional AuNP assay (Fig. 4c, Supplementary Information Section 7), an increase of 43 percentage points in the total patients. Real-world testing involves individuals at various stages of infection, so we also estimate the impact at the population level on a single testing day using the respective assay sensitivity across all viral loads, independent of individual infection stages. This analysis suggests that, at the peak of the Omicron wave in the U.K. (Jan 2022) the sensitivity of the FND assay is estimated to detect up to 69,400 more patients in single day than our conventional AuNP assay (Supplementary Information Section 8, Supplementary Information Fig. 4). This approach provides an estimate of the number of additional patients that could be detected in a population-wide setting given improved sensitivity of the FND assay.

**Fig 4:**
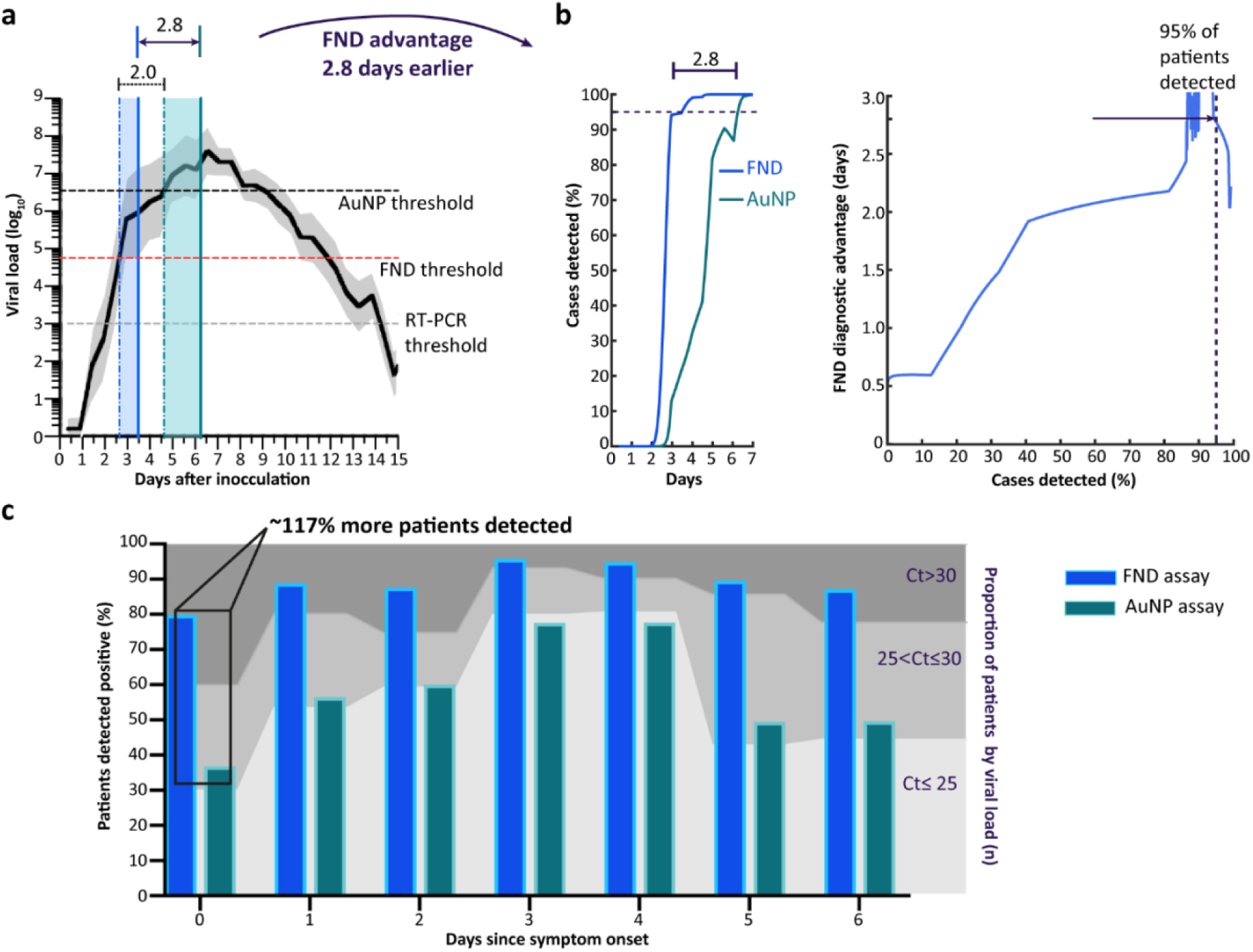
Modelling diagnostic advantage of FND assay compared to AuNPs. **(a)** Viral load infection dynamics adapted from Killingley et al.^25^ human challenge trial tracking viral load over the first 15 days of infection. Black line tracks mean patient data with grey region indicating 95% credible interval, using combined uncertainty from the assay thresholds and distribution of patient viral loads. The horizontal dashed lines represent assay thresholds at 10^3^ copies/mL for RT-PCR (grey), 5.6×10^4^ copies/mL for FND (red) and 3.5×10^6^ copies/mL for AuNP black). The mean day of initial detection was 2.6 days [95% credible interval: 2.6-2.7] with the FND assay vertical blue dashed line), and ∼4.6 days [95% credible interval: 4.4-4.8] with AuNPs (vertical green dashed line). Taking the difference in means days shows that the FND assay can 2 days earlier [95% credible interval: 1.8-2.1], on average (p-value< 0.0001, t = 25). The shaded regions represent the gap between the detection of 50% of patients (vertical dashed lines) and 95% of patients with 95% confidence (vertical solid lines). We also show the FND assay could detect patients within 0.6 days of RT-PCR **(b)** The FND assay will detect infection in 95% (blue solid line) of patients no later than 3.5 days compared to the AuNP assay ≤ 6.3 days, a 2.8-day diagnostic advantage for detection with FNDs. (c) Line graph shows data adapted from Frediani et al.,^48^ representing the proportion of patient samples within ranges Ct≤25, 25<Ct≤30, and Ct>30 on each day from symptom onset (day 0). The bar graph shows the sensitivity of each assay weighted by proportion of high, moderate, and low viral load samples detected each day. Compared to the AuNP assay, the FND assay could detect ∼117% (2.2-fold) more patients on the first day of infection (symptom onset) and ∼57% (1.6-fold) more patients the second day.

This work demonstrates a high level of concordance with RT-qPCR. However, interpretation of antigen sensitivity in low viral load samples using RT-qPCR Ct cut-offs should be taken with caution as (i) Ct values vary depending on the assay^50,51^, and (ii) RT-qPCR can suffer from extended RNA positivity where thresholds for infectiousness in relation to Ct values has yet to be fully established^52^. Notably, Killingley et al.^25^, reported quantifiable virus by RT-qPCR was still present by day 14 necessitating extended quarantine. In contrast, viable virus by cell culture, often used as a better indicator of infectiousness, showed viral clearance by day 10.2 on average and no later than day 12. This supports compounding evidence that Ag-LFTs are good determinants of infectiousness, and more closely aligns with the FND assay thresholds where our fitting shows latest detection at day 12 (Fig. 4a). Therefore, 100% sensitivity across the whole range of RT-qPCR Ct values may not be desired. The reduction in sensitivity in low viral load samples (<10^4^ copies/mL) may be beneficial for differentiating active viral infection and residual circulating antigen. Further standardisation is needed on SARS-CoV-2 antigen test sensitivity and the minimum viral load needed to determine infectiousness.

Interpretation of sensitivity in SARS-CoV-2 clinical studies depends on sample selection and assay thresholds (e.g. evaluations including sample pools skewed to high viral loads can cause overestimation of test sensitivity than would be achieved in the general population). The initial random selection of samples in this study, from hospitalised patients to travellers, is somewhat representative of samples found in the general population. The second set of samples targeting Ct>25 may skew the sample pool to the low viral end, suggesting an underestimate of overall clinical sensitivity. In addition, although this work did not find the swab type (nasopharyngeal and nasal only) had a significant impact on device performance (Supplementary information Fig. 3), variability in viral load and resulting Ag-LFT performance have been reported in literature^25,53,54^. Further standardisation is needed to determine prevalence within these grouped Ct sample pools and swab type should be consistent so that pre-clinical device performance evaluations accurately depict performance in the field. Accurate determination of thresholds set by the negative samples can also impact test sensitivity and specificity. Our negative sample size of n=50 gives a standard error of the mean of ±14% of σ, where σ is the true standard deviation of negative signal values, and standard error of σ of ±10% of σ. This is sufficiently small to accurately estimate the threshold. The FNDs high sensitivity also presents a promising avenue for early diagnosis in other diseases, like influenza where early administration of Tamiflu within 48 hours of symptom onset can significantly improve health outcomes^55^, and future paradigms such as HIV viral load self-monitoring on a LFTs^56^.

Future work involves moving towards a cassette-based lateral flow test platform with integrated reagents. Initial experiments described in Supplementary Information section 9 demonstrate adequate reaction kinetics required for an integrated device where capture and detection of reagents typically occurs during rehydration of the conjugate and flow up the membrane (Supplementary information Fig. 5). The lateral flow strip read-out is a key consideration for fluorescence-based assays. More specific to our FND-based modulated platform, we considered the dielectric effect from a wet lateral flow strip on the resonator and subsequent lock-in result to allow for immediate read-out. The effect of wetting of the lateral flow showed a ∼40% reduction (small compared to the LOD uncertainty and comparable to the strip-to-strip coefficient of variation of 18.3%) in the lock-in signal with immediate read-out after LFT running compared to after 20 minutes (Supplementary Information Fig. 6). However, this effect can be accounted for by normalising to the test membrane, or control line, or using a tuneable resonator to mitigate shifts in resonant frequency as the strip dries (Supplementary Information section 9b). The small size of the resonator and high brightness of FNDs allow for this system to be integrated into a portable, cost-effective smartphone fluorescent reader with estimated production cost of our prototype device at ∼£912 (Supplementary Information section 9c, Supplementary Information Table 6).

To conclude, we demonstrate that spin-enhanced FNDs are a highly sensitive, specific and versatile platform for antigen detection, achieving a 1,100-fold improvement over gold nanoparticles using identical capture antibodies, giving a 2.8-day diagnostic advantage to detect virus in 95% of patients, and could have detected up to 69,400 more cases at the peak of the Omicron wave. Spin-enhanced FNDs showed 95.1% sensitivity in clinical samples (Ct≤30) and 100% specificity compared to RT-qPCR. The sensitivity achieved is comparable to high end Quanterix biomarker discovery platforms at a fraction of the cost. Our findings demonstrated the utility of nanodiamonds in a large blinded clinical study, laying the foundations for future research to explore antigen detection in clinical samples with high background and assays with low nonspecific binding. Future research is needed to miniaturisation of optical and modulation components into a portable fluorescent reader to improve access for low-resource and point of care settings, for example A&E, pharmacies, and home monitoring (e.g. disease or drug). Additionally, a larger clinical study is needed, and extension to other diseases, both communicable and non-communicable.

## Methods

### Biolayer Interferometry for antibody binding kinetics

Biolayer interferometry (Fortebio Octet Red) was used to analyse the antibody binding kinetic parameters. The assay was performed using a streptavidin-coated biosensor (Octet® Streptavidin (SA) biosensor, P/N 18-5019). A volume of 200μL sample or kinetics buffer (P/N 18-5032) was added to a 96-well plate (Greiner bio-one) for each baseline, loading, association, and dissociation step. Biotinylated nucleocapsid protein (Sino biological Cat# 40588-V08B-B) was loaded on the SA sensor at 1μg/mL. Following a baseline step, the biosensors moved to wells containing a 2-fold dilution series of the target nucleocapsid antibody (CR3009, CR3018, 40143-R001, 40143-R004, 40143-R040, 40143-MM08) diluted in kinetic buffer for the association step. Antibody concentrations ranged from 20 μg/mL to 0.001 μg/mL with each sample performed in triplicate. The binding curves were analysed in MATLAB using a 1:1 binding kinetics model.

### Fluorescent nanodiamond functionalisation

Polyglycerol (GF)-coated FNDs 600nm were conjugated to antibodies using disuccinimidyl carbonate (DSC) following conjugation protocol in Miller et al. Antibody functionalisation concentration was adapted for 600nm particles considering surface area to volume ratio where 2.71μg of antibody (1mg/mL) was added to 100μL of particles at 1 mg/mL stock concentration. The functionalised FND concentration was then measured by fluorescent intensity against a standard curve as described in Miller et al.

### Particle characterisation

Dynamic light scattering (Zetasiser Nanoseries, Malvern Instruments Ltd) was used to characterise the hydrodynamic diameter and monodispersity-polydispersity index (PDI) of the nanoparticles. Samples for scanning electron microscopy (SEM) were prepared by drop casting the 600 FND-PG in 12.5mM MgCl_2_ in PBS and dried for 30mins. The addition of 12.5mM MgCl_2_ improves particle adhesion to the substrate surface. Samples were imaged on the focused ion beam Zeiss XB1540 microscopy system. Quantitative analysis of particle size was performed across 5 SEM images with a total count of 156 FNDs (ImageJ). The width and diagonal (longest dimension) of each particle were measured with reference to the SEM scale bar. These measurements were then used to calculate particle size, assuming an approximate spherical shape. The 600nm FND excitation spectrum was measured on a plate reader (Clariostar/Molecular devices) by sweeping excitation wavelengths from 320nm to 640nm with the emission filter set to 675nm. The emission spectrum was measured by a widefield fluorescent microscope coupled to an Optosky ATP2000P-340-850-100 spectrometer at 2ms integration time.

### FND lateral flow testing

The lateral flow assays were ran using commercially available strips with polystreptavidin printed test lines (Global Access Diagnostics). FND-PG-AbMM08 was diluted in PBS to concentration of 26.4fM. For the direct bind format, 5μL of the FNDs + 50μL of biotinylated nucleocapsid protein diluted in running buffer (150mM Tri-HCL pH 8, 2% IGEPAL CA-630, 100mM NaCl, 0.05%Tween20, 0.8%Casein) was added to wells in a 96-well plate and allowed to bind for 10mins. The strips were added to each well and allowed to run for ∼15 mins. Strips were allowed to dry before read-out described in the following section. Full microscope set up is described in Miller et al^15^. For the sandwich format assay, FND-PG-AbMM08 was diluted in PBS to concentration of 26.4 fM and secondary biotinylated AbR001 was diluted in PBS to 513nM. 5μL of the FNDs + 49μL of sample (nucleocapsid protein or gamma-irradiated virus) diluted in running buffer + 1μL of bAb01 (513nM) was added to wells in a 96-well plate and allowed to bind for 10mins. Strip were added to the wells and run for 15 mins, followed by test line read-out of dry strips.

### FND test line analysis

The lateral flow strips were imaged using a fluorescence microscope (Olympus BX51) with a 550nm green LED excitation light source (CoolLED pE-4000), a filter cube with an excitation filter (Semrock-500nm bandpass, 49nm bandwidth), a dichroic mirror (Semrock-596nm edge), and 593nm long-pass emission filter (Semrock). A 20x/0.4 BD objective lens was used. Images were recorded using a high-speed camera (Hamamatsu, ORCA-Flash4.0 V3) and HCImage Live software (Hamamatsu) where the mean of each frame were calculated to give a time-series of mean pixel values. A voltage controlled oscillator (VCO) (Mini-Circuits-ZX95-3360+) and an amplifier (Mini-Circuits-ZX60-33LN+) were connected to an antenna resonator and circuit board (Minitron, Rogers 4003c 0.8mm substrate with 300gm-2 copper weight) to generate the microwave field. The resonator was designed to match the frequency of ∼2.87GHz to produce desirable contrast of FNDs at the test line (Miller et al.^15^). The modulated signal is achieved by modulating the VCO input with a reference frequency generator at 4Hz, using a 32.768Hz crystal oscillator (Farnell, DS32KHZ) and 14-stage frequency divider (Farnell, CD4060BM). The microwave circuit was designed by B. Miller in previous work. The fluorescence signal was modulated with a set modulation frequency (*Fm*) and the amplitude of the modulating signal was processed using a computational lock-in algorithm via MATLAB. This computational lock-in algorithm is described in Miller et al.^15^

### Optically detected magnetic resonance (ODMR) measurements

An ODMR measurement was taken from a strong positive test line on the nitrocellulose strip to validate the fluorescence output from the FND NV^-^ centres in the presence of a microwave field. A lateral flow strip with 600nm FND-PG-AbMM08 bound to the test line in a direct bind format via biotinylated nucleocapsid protein (10ng/mL) was placed under the microscope set-up with microwaves being supplied using a (SynthUSB3) source through a stripline and a high-power amplifier (12V, ZRL-3500+, Mini Circuits) to sweep across frequencies from 1 to 3 GHz. A MATLAB script was used to plot the frequency intensity across different frequencies. Limit-of-detection analysis A 2-fold serial dilution of recombinant nucleocapsid protein or SARS-CoV-2 gamma irradiated virus was ran on the half-strip dipstick sandwich format assay and the lock-in intensity values were used to determine the limit of detection. The LoD was plotted and calculated based on a Langmuir model fit using a MATLAB software tool developed by Miller et al.^42^, which is available open source on Github^57^. A statistical method for determining the limit of detection (LoD) was employed based on methods reported in Holstein et al., and adapted by Miller et al.^42^

### Evaluation of clinical swab samples

Nasal and nasopharyngeal swab samples were collected from individuals at UCLH (including travelers, healthcare workers, or other patients) from Nov-Dec 2022 and June-Aug 2023. Ct values were initially determined following HSL in-house standard protocol. Residual samples were stored at -80°C until further testing. Samples were randomly selected by UCLH for transfer and tested blinded on the FND Ag-LFT in-house. Subsequently, samples were unblinded for analysis and positive samples were re-evaluated in-house by RT-qPCR using N1 2019-nCoV RUO kit (Integrated DNA Technologies Cat#10006713) on QuantStudio RT-qPCR system (ThermoFisher Scientific). 5-point 10-fold dilution series of SARS-CoV-2 synthetic RNA positive control (Twist Bioscience, control 51 cat#105346) was used for standard curve quantification to RNA copies/mL. For lateral flow testing, 40μL of swab sample in VTM was added to the well and mixed with 9μL of 5X running buffer (see Supplementary Information section 1b for buffer formulation) + 5μL of the FNDs (26 fM) + 1μL of AbR001-b (513nM) and allowed to bind for 10mins. Strips were added to wells and ran for 15mins. The strips were allowed to dry before read-out described in the previous section.

### Inclusion & Ethics

The UCLH Governance Committee has approved the study performed under the UCLH HTA license for use of residual samples for development of diagnostic assays (NDU-VIR_131/13122022). All samples where pseudo-anonymised at source and tested prior to discard. Sex, gender, race, or ethnicity information were not disclosed to or considered by investigators in this study.

## Data Availability

All data produced in the present study are available upon reasonable request to the authors

## Acknowledgments

This work was funded by the i-sense EPSRC IRC in Agile Early Warning Sensing Systems in Infectious Diseases and Antimicrobial Resistance (EP/R00529X/1); the National Institute for Health Research University College London Hospitals Biomedical Research Centre; the EPSRC Digital Health Hub for AMR (EP/X031276/1); a London Centre for Nanotechnology Departmental Studentship to A.T.D., Wellcome Trust 224071/Z/21/Z to B.S.M. and the UCLH NHS Foundation Trust to J.B. and E.N. We thank Peter Cherepanov (Francis Crick Institute) for providing recombinant nucleocapsid protein and Laura McCoy (UCL) for providing antibodies CR3009 and CR3018.

## Contributions

A.T.D., B.S.M and R.A.M. conceived the research and led the study; A.T.D. designed and executed all experiments; B.S.M. co-supervised A. T. D., and the analysis of Forte bio kinetic data, LFT LOD analysis and statistics using bespoke software, and modelling clinical threshold Bayesian analysis; D.H. co-supervised A.T.D. and helped with the RT-qPCR experiment and analysis; F.D. and M. M. assisted with nanodiamond optical characterisation; E.N. and J.B. provided clinical expertise and clinical samples; C.K.O provided recombinant protein; L.E.M provided antibodies; A.T.D., B.S.M, F.D and R.A.M. drafted the manuscript; and all authors reviewed and revised the manuscript.

## Competing interests

B.S.M and R.A.M are inventors on the UK patent application number 1814532.6 filed by University College London Business.

## Data availability

The data that support the findings of this study are available from the corresponding authors, R.A.M and B.S.M, upon reasonable request. Source data will be made available with this paper.

## Code availability

The MATLAB software for fitting robust detection limits (LODs) and confidence intervals to serial dilution data is available at https://github.com/bensmiller/detection-limit-fitting/. The code used for binding kinetics and Bayesian analysis are available from the corresponding authors, R.A.M and B.S.M, upon reasonable request.

---

**Extended Data Fig. 1:**
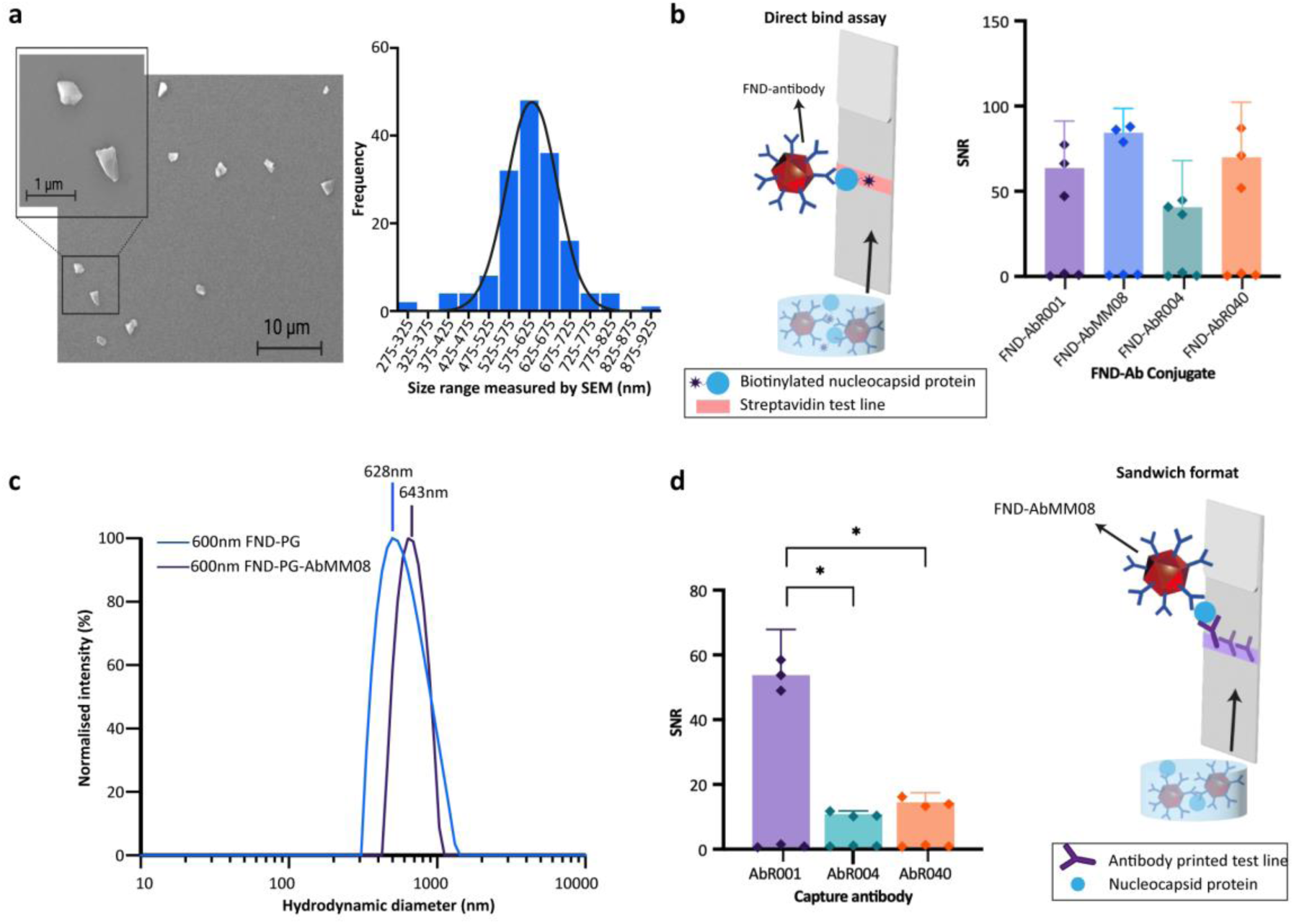
FND characterisation and antibody pair selection. **(a)** Scanning electron microscopy (SEM) image (left) of polyglycerol-coated 600nm FNDs at magnification of 3.21 K X showing monodispersed particles. Call-out box shows particles at magnification 27.03 K X showing cubic morphology of particles. Histogram (right) with size dispersion of the 600nm FNDs based on counts from SEM scans, showing distribution across size ranges from 275nm to 925nm, with peak counts between 575-625nm particles of size 575-625nm. Distribution is fit to a gaussian curve (black line) with mean=606.9 nm (± 61.7) **(b)** Schematic of direct bind assay and plot of mean signal-to-noise ratio (n=3) with standard error of means using different antibodies functionalised to 600nm FNDs with FND-AbMM08 showing best SNR of 83, but with no statistically significant difference (p-value=0.71, ANOVA, F=0.48, DF=11, post-hoc Tukey’s multiple comparisons). Data points represent the SNR of individual test replicates using the mean baseline signal (n=3). **(c)** Mean FND size distribution plot using dynamic light scattering showing particle intensity distribution (%) of both 600nm-FND-PG and antibody functionalised 600nm-FND-PG with hydrodynamic diameters of 628nm (±220), and 643nm (±140).(d) SNR of different antibody-printed test lines (Supplementary Information section 1c) with FND-AbMM08 conjugate. Positive sample was 10ng/mL recombinant protein (n=3). Error bars represent standard error of mean. Data points represent the SNR of individual test replicates replicates using the mean baseline signal (n=3). Statistical significance determined by one-way ANOVA, F=8.2, DF=8, post-hoc Tukey’s multiple comparisons, * denotes p-values <0.05: 0.0251 and 0.0363 for AbR001 vs. AbR004 and AbR040, respectively. No significant difference observed between AbR004 and AbR040 (p-value=0.95).

**Extended Data Fig. 2:**
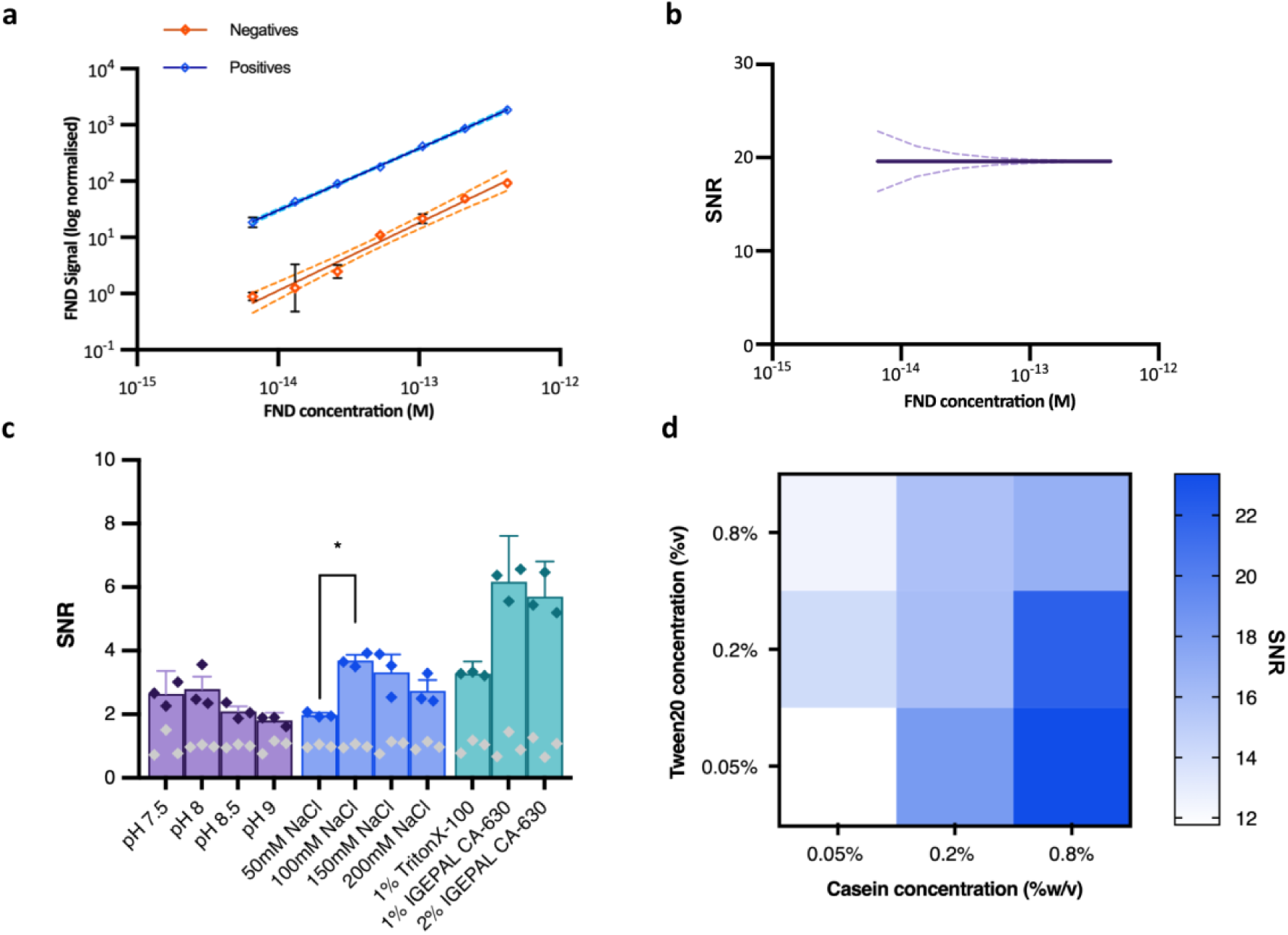
FND Ag-LFT optimisation. **(a)** FND concentration sweep from 6.6 fM to 42 pM tested on a negative sample (n=3) and positive sample (500pg/mL recombinant nucleocapsid protein n=3). Data points show mean and error bars show standard deviation (error bars not shown represent s.d < 0.05 units). Plots are fitted with linear regressions in Graphpad Prism; negatives (orange) R-squared=0.9545 and positives (blue) R-squared=0.9978. **(b)** SNR found by dividing the fitted linear regressions in **(a)**, resulting in a constant value of ∼20. Dashed line represents s.d. error. **(c)** SNR testing different buffer components for viral lysis using whole inactivated virus sample (7.9 x 10^3^ TCID_50_/mL) as positive sample (n=3). Buffer pH8 was selected and tested with addition of NaCl, where 100 mM NaCl only showed significant difference from 50 mM (p-value=0.0323, one-way ANOVA, F=4.70,n=8, post-hoc Tukeys multiple comparisons). IGEPAL CA-630 showed better SNR at both concentrations although not a statistically significant difference determined by one-way ANOVA p-value=0.21, F=2.1, n=9, post-hoc Tuckey’s multiple comparisons. Error bars represent standard error of the mean. Data points represent the SNR of individual test replicates by mean baseline signal. Buffer was tested independent of additional protein blockers (shown in part d) resulting in overall poorer contrast **(d)** Varying concentrations of Tween20 and Casein in the buffer to determine optimal concentration of blocking components. Heat map shows mean SNR of a negative (n=3) and positive (500pg/mL recombinant nucleocapsid protein n=3) where the addition of 0.8% casein and 0.05% Tween20 showed the best SNR of 23.

**Extended Data Fig. 3:**
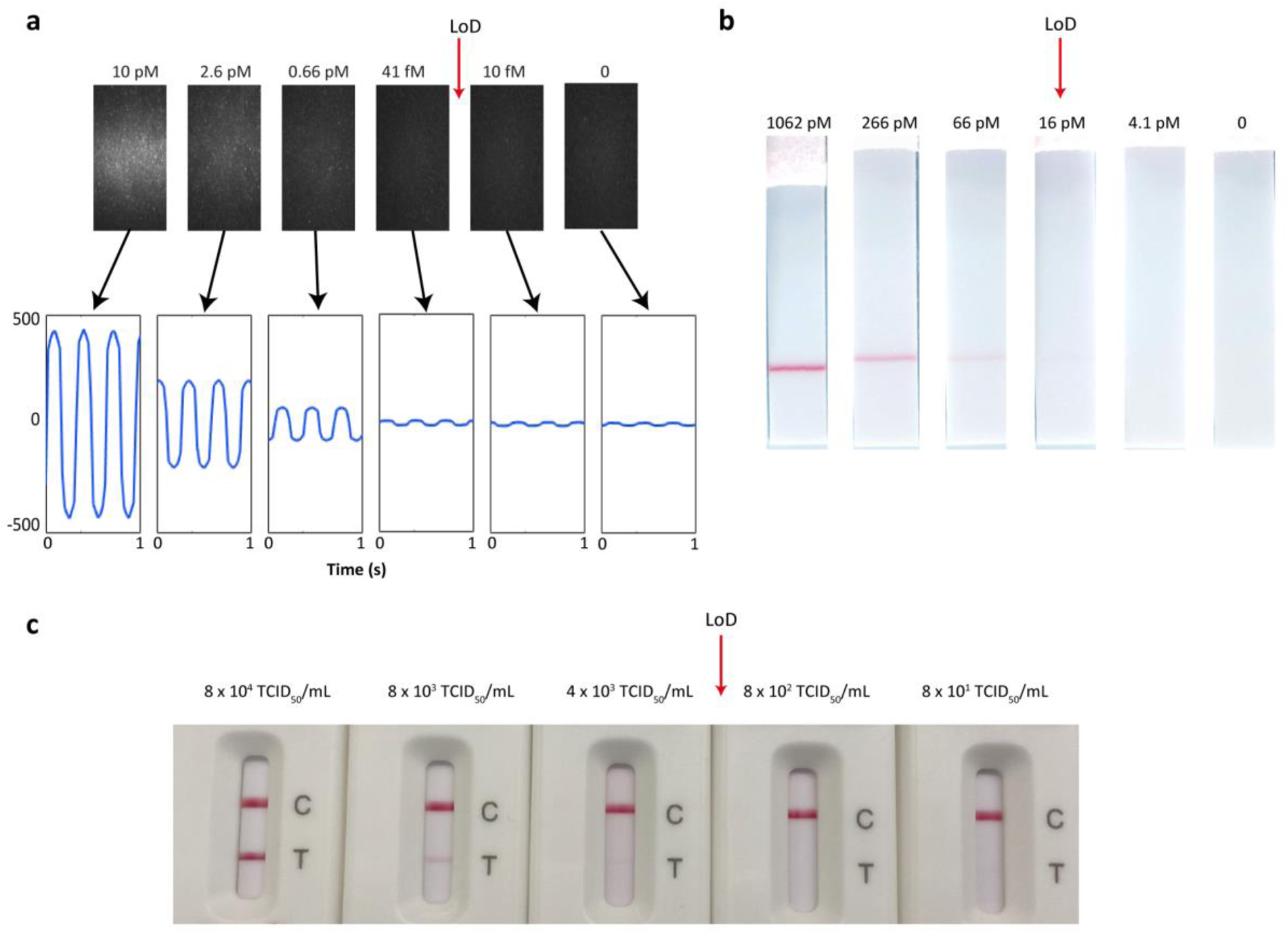
Assay test line analysis across FND LFT, in-house gold LFT and a commercial test. **(a)** LoD with recombinant nucleocapsid protein on FND assay. Images of FND Ag-LFT test lines from selected concentrations of serial dilution with corresponding fluorescent intensity time series plots (bottom) showing detectable periodic signal after test line is no longer visible in the images. Arrow shows the approximate LoD calculated from the exponential fit (Fig. 2b). **(b)** Select concentrations of strips from serial dilution of recombinant nucleocapsid protein. Samples were ran in triplicate for each sample concentration. AuNP assay strips were ran for 15mins and imaged by camera in a light-controlled box. Images of the test line were then analysed through MATLAB using a pixel-wise line intensity plot. Arrow points to the calculated LoD from exponential fitting curve. **(c)** Images of FlowFlex SARS-CoV-2 Ag assay tested with selected concentrations of gamma irradiated wild-type viral isolate following the product insert protocol. Arrow indicates approximate concentration of lowest detectable test line at ∼10^3^ TCID_50_/mL.

**Extended Data Fig. 4:**
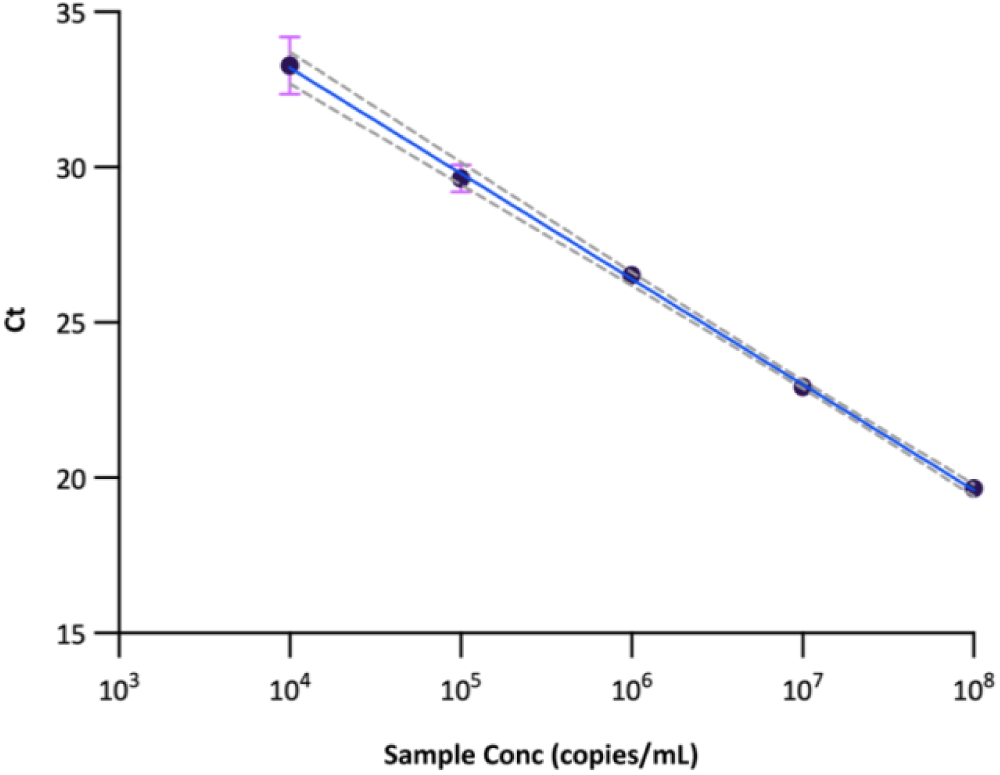
RT-qPCR standard curve. RT-qPCR standard curve of standard Omicron viral RNA (n=3) to convert Ct to copies/mL. The data were fitted to a Bayesian linear regression weighted by the uncertainty in Ct. The fit, along with error propagation was used to calculate copies/mL and associated errors from Ct values from clinical samples. Errors were calculated by combining the uncertainty in the model (sampling from the posterior to get the distribution of copies/mL values for a given Ct value), and the uncertainty in the measured Ct value (from interpolating the relationship between Ct and Ct standard error).

**Extended Data Fig. 5:**
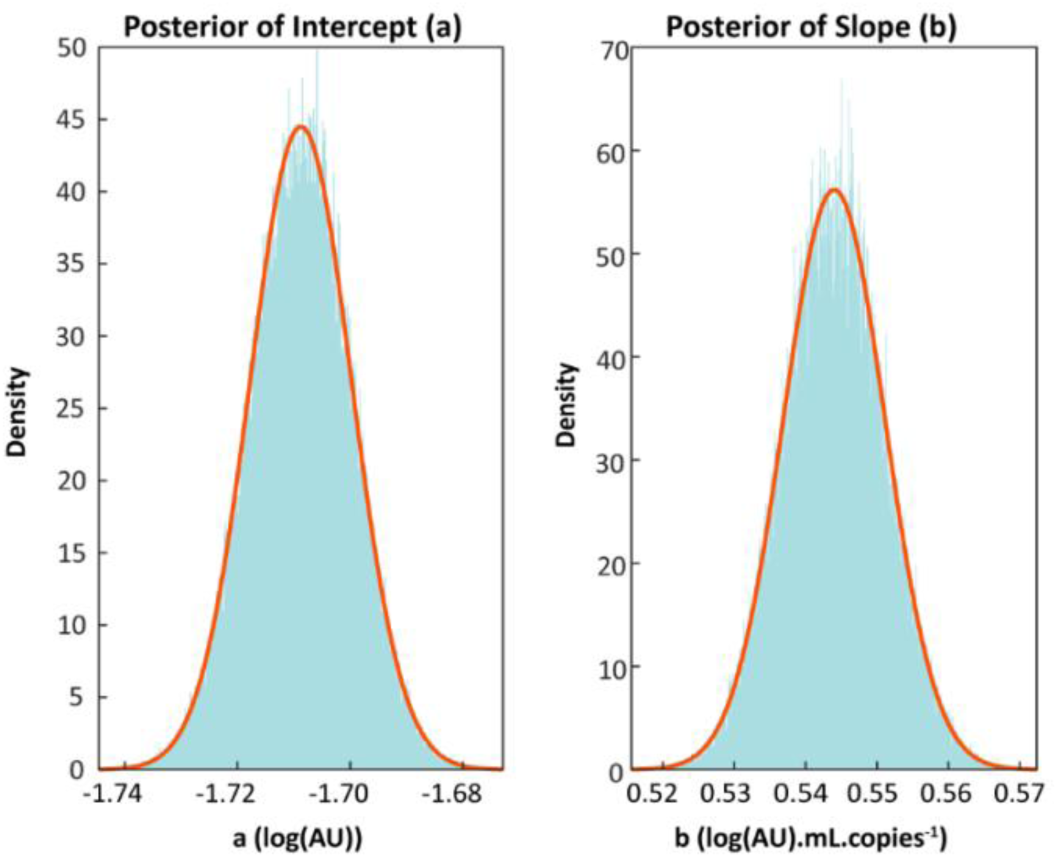
Clinical sample Bayesian regression posterior distributions. Posterior distributions for the intercept and slope for the symmetrical Bayesian linear regression with errors-in-variables in x and y shown in Fig. 3b. See Supplementary Information Section 6 for priors and likelihood functions. Blue histograms show Markov chain Monte Carlo sampling of the posteriors and the red lines show Gaussian distributions with means and standard deviations calculated from the distributions.

## Notes

### Author Declarations

The University College London Hospital Governance Committee gave ethical approval for this study performed under the University College London Hospital Human Tissue Act license for use of residual samples for development of diagnostic assays (NDU-VIR_131/13122022). All samples where pseudo-anonymised at source and tested prior to discard.

## References

1. Aslam, N. et al. Quantum sensors for biomedical applications. Nature Reviews Physics 5, 157–169 (2023).

2. Doherty, M. W. et al. The nitrogen-vacancy colour centre in diamond. Phys Rep 528, 1–45 (2013).

3. Bucher, D. B. et al. Quantum diamond spectrometer for nanoscale NMR and ESR spectroscopy. Nat Protoc 14, 2707–2747 (2019).

4. McGuinness, L. P. et al. Quantum measurement and orientation tracking of fluorescent nanodiamonds inside living cells. Nat Nanotechnol 6, 358–363 (2011).

5. Gu, Q. et al. Simultaneous Nanorheometry and Nanothermometry Using Intracellular Diamond Quantum Sensors. ACS Nano 17, 20034–20042 (2023).

6. Glenn, D. R. et al. Single-cell magnetic imaging using a quantum diamond microscope. Nat Methods 12, 736–738 (2015).

7. Atallah, J. et al. Rapid Quantum Magnetic IL-6 Point-of-Care Assay in Patients Hospitalized with COVID-19. Diagnostics vol. 12 Preprint at 10.3390/diagnostics12051164 (2022).

8. Chen, S. et al. Digital Magnetic Detection of Biomolecular Interactions with Single Nanoparticles. Nano Lett 23, 2636–2643 (2023).

9. Fan, S., Lopez Llorens, L., Perona Martinez, F. P. & Schirhagl, R. Quantum Sensing of Free Radical Generation in Mitochondria of Human Keratinocytes during UVB Exposure. ACS Sens 9, 2440–2446 (2024).

10. Paterson, A. S. et al. Persistent luminescence strontium aluminate nanoparticles as reporters in lateral flow assays. Anal Chem 86, 9481–9488 (2014).

11. Reineck, P. et al. Brightness and Photostability of Emerging Red and Near-IR Fluorescent Nanomaterials for Bioimaging. Adv Opt Mater 4, 1549–1557 (2016).

12. Mochalin, V. N., Shenderova, O., Ho, D. & Gogotsi, Y. The properties and applications of nanodiamonds. Nat Nanotechnol 7, 11–23 (2012).

13. Igarashi, R. et al. Real-Time Background-Free Selective Imaging of Fluorescent Nanodiamonds in Vivo. Nano Lett 12, 5726–5732 (2012).

14. Shah, K. G. & Yager, P. Wavelengths and Lifetimes of Paper Autofluorescence: A Simple Substrate Screening Process to Enhance the Sensitivity of Fluorescence-Based Assays in Paper. Anal Chem 89, 12023–12029 (2017).

15. Miller, B. S. et al. Spin-enhanced nanodiamond biosensing for ultrasensitive diagnostics. Nature 587, 588–593 (2020).

16. Wei-Wen Hsiao, W., et al. Fluorescent nanodiamond-based spin-enhanced lateral flow immunoassay for detection of SARS-CoV-2 nucleocapsid protein and spike protein from different variants. Anal Chim Acta 1230, 340389 (2022).

17. Le, T. N. et al. Fluorescent nanodiamond immunosensors for clinical diagnostics of tuberculosis. J Mater Chem B 12, 3533–3542 (2024).

18. Feuerstein, G. Z. et al. The Use of Near-Infrared Light-Emitting Fluorescent Nanodiamond Particles to Detect Ebola Virus Glycoprotein: Technology Development and Proof of Principle. Int J Nanomedicine 15, 7583–7599 (2020).

19. WHO. An R&D Blueprint for Action to Prevent Epidemics. https://www.who.int/publications/m/item/an-r-d-blueprint-for-action-to-prevent-epidemics (2016).

20. Hu, B., Guo, H., Zhou, P. & Shi, Z.-L. Characteristics of SARS-CoV-2 and COVID-19. Nat Rev Microbiol 19, 141–154 (2021).

21. WHO. WHO Director-General’s Opening Remarks at the Media Briefing on COVID-19–16 March 2020. World Health Organization 4 Preprint at https://www.who.int/director-general/speeches/detail/who-director-general-s-opening-remarks-at-the-media-briefing-on-covid-1916-march-2020 (2020).

22. WHO. Laboratory testing for 2019 novel coronavirus (2019-nCoV) in suspected human cases. WHO - Interim guidance vol. 2019 1–7 Preprint at http://files/2702/World Health Organization - 2020 - Laboratory testing for 2019 novel coronavirus (2019-nCoV) in suspected human cases.pdf (2020).

23. Budd, J. et al. Lateral flow test engineering and lessons learned from COVID-19. Nature Reviews Bioengineering 1, 13–31 (2023).

24. Peeling, R. W., Olliaro, P. L., Boeras, D. I. & Fongwen, N. Scaling up COVID-19 rapid antigen tests: promises and challenges. Lancet Infect Dis 3099, 21–26 (2021).

25. Killingley, B. et al. Safety, tolerability and viral kinetics during SARS-CoV-2 human challenge in young adults. Nat Med 28, 1031–1041 (2022).

26. Peeling, R. W., Heymann, D. L., Teo, Y.-Y. & Garcia, P. J. Diagnostics for COVID-19: moving from pandemic response to control. The Lancet 399, 757–768 (2022).

27. Mina, M. J., Parker, R. & Larremore, D. B. Rethinking Covid-19 Test Sensitivity — A Strategy for Containment. New England Journal of Medicine 383, e120 (2020).

28. Puhach, O., Meyer, B. & Eckerle, I. SARS-CoV-2 viral load and shedding kinetics. Nat Rev Microbiol 21, 147–161 (2023).

29. Pickering, S. et al. Comparative performance of SARS-CoV-2 lateral flow antigen tests and association with detection of infectious virus in clinical specimens: a single-centre laboratory evaluation study. Lancet Microbe 2, e461–e471 (2021).

30. Brümmer, L. E., et al. Accuracy of Novel Antigen Rapid Diagnostics for SARS-CoV-2: A Living Systematic Review and Meta-Analysis. PLoS Medicine vol. 18 (2021).

31. Wagenhäuser, I. et al. Clinical performance evaluation of SARS-CoV-2 rapid antigen testing in point of care usage in comparison to RT-qPCR. EBioMedicine 69, (2021).

32. Scheiblauer, H. et al. Comparative sensitivity evaluation for 122 CE-marked rapid diagnostic tests for SARS-CoV-2 antigen, Germany, September 2020 to April 2021. Eurosurveillance 26, (2021).

33. Schrand, A. M., Hens, S. A. C. & Shenderova, O. A. Nanodiamond Particles: Properties and Perspectives for Bioapplications. Critical Reviews in Solid State and Materials Sciences 34, 18–74 (2009).

34. Liu, Y., Zhan, L., Qin, Z., Sackrison, J. & Bischof, J. C. Ultrasensitive and Highly Specific Lateral Flow Assays for Point-of-Care Diagnosis. ACS Nano 15, 3593–3611 (2021).

35. Yetisen, A. K., Akram, M. S. & Lowe, C. R. Paper-based microfluidic point-of-care diagnostic devices. Lab Chip 13, 2210–2251 (2013).

36. Gasperino, D., Baughman, T., Hsieh, H. V., Bell, D. & Weigl, B. H. Improving Lateral Flow Assay Performance Using Computational Modeling. Annual Review of Analytical Chemistry 11, 219–244 (2018).

37. Cate, D. M. et al. Antibody Screening Results for Anti-Nucleocapsid Antibodies Toward the Development of a Lateral Flow Assay to Detect SARS-CoV-2 Nucleocapsid Protein. (2021) doi:10.1021/acsomega.1c01253.

38. Grant, B. D. et al. A SARS-CoV-2 coronavirus nucleocapsid protein antigen-detecting lateral flow assay. PLoS One 16, e0258819 (2021).

39. Bachman, C. M. et al. Clinical validation of an open-access SARS-COV-2 antigen detection lateral flow assay, compared to commercially available assays. PLoS One 16, e0256352 (2021).

40. Olsen, D. A. et al. Quantifying SARS-CoV-2 nucleocapsid antigen in oropharyngeal swabs using single molecule array technology. Sci Rep 11, 20323 (2021).

41. van den Brink, E. N., et al. Molecular and biological characterization of human monoclonal antibodies binding to the spike and nucleocapsid proteins of severe acute respiratory syndrome coronavirus. J Virol 79, 1635–1644 (2005).

42. Miller, B. S. et al. Sub-picomolar lateral flow antigen detection with two-wavelength imaging of composite nanoparticles. Biosens Bioelectron 207, 956–5663 (2022).

43. Wang, C. et al. Ultrasensitive and Simultaneous Detection of Two Specific SARS-CoV-2 Antigens in Human Specimens Using Direct/Enrichment Dual-Mode Fluorescence Lateral Flow Immunoassay. ACS Appl Mater Interfaces 13, 40342– 40353 (2021).

44. Chen, W. et al. An integrated fluorescent lateral flow assay for multiplex point-of-care detection of four respiratory viruses. Anal Biochem 659, 114948 (2022).

45. Gupta, R. et al. Ultrasensitive lateral-flow assays via plasmonically active antibody-conjugated fluorescent nanoparticles. Nat Biomed Eng 7, 1556–1570 (2023).

46. Cubuk, J. et al. The SARS-CoV-2 nucleocapsid protein is dynamic, disordered, and phase separates with RNA. Nat Commun 12, 1–17 (2021).

47. Dinnes, J. et al. Rapid, point-of-care antigen tests for diagnosis of SARS-CoV-2 infection. Cochrane Database of Systematic Reviews 2022, (2022).

48. Frediani, J. K. et al. The New Normal: Delayed Peak SARS-CoV-2 Viral Loads Relative to Symptom Onset and Implications for COVID-19 Testing Programs. Clinical Infectious Diseases 78, 301–307 (2024).

49. Alexander, V. W. et al. Daily SARS-CoV-2 Nasal Antigen Tests Miss Infected and Presumably Infectious People Due to Viral Load Differences among Specimen Types. Microbiol Spectr 11, e01295–23 (2023).

50. Walker, A. S. et al. Ct threshold values, a proxy for viral load in community SARS-CoV-2 cases, demonstrate wide variation across populations and over time. Elife 10, e64683 (2021).

51. Rhoads, D. et al. College of American Pathologists (CAP) Microbiology Committee Perspective: Caution Must Be Used in Interpreting the Cycle Threshold (Ct) Value. Clinical Infectious Diseases 72, e685–e686 (2021).

52. Cevik, M. et al. SARS-CoV-2, SARS-CoV, and MERS-CoV viral load dynamics, duration of viral shedding, and infectiousness: a systematic review and meta-analysis. Lancet Microbe 2, e13–e22 (2021).

53. Glenn, P. et al. Comparison between Nasal and Nasopharyngeal Swabs for SARS-CoV-2 Rapid Antigen Detection in an Asymptomatic Population, and Direct Confirmation by RT-PCR from the Residual Buffer. Microbiol Spectr 10, e02455–21 (2022).

54. Brümmer, L. E. et al. Accuracy of novel antigen rapid diagnostics for SARS-CoV-2: A living systematic review and meta-analysis. PLoS Med 18, e1003735-(2021).

55. Aoki, F. Y. et al. Early administration of oral oseltamivir increases the benefits of influenza treatment. Journal of Antimicrobial Chemotherapy 51, 123–129 (2003).

56. K, D. P., et al. Point-of-Care HIV Viral Load Testing: an Essential Tool for a Sustainable Global HIV/AIDS Response. Clin Microbiol Rev 32, 10.1128/cmr.00097-18 (2019).

57. Miller, B. S. Detection Limit Fitting Tool. GitHub https://github.com/bensmiller/detection-limit-fitting (2022).

58. Dellaportas, P. & Stephens, D. A. Bayesian Analysis of Errors-in-Variables Regression Models. Biometrics 51, 1085–1095 (1995).

