## Supplementary Information for "Quantum-enhanced nanodiamond rapid test advances early SARS-CoV-2 antigen detection in clinical diagnostics"

### 1) Supplementary methods

#### a) Materials and reagents

FortéBio Octet Red96 Streptavidin sensor plates (Part No 18-5019) and Kinetics buffer 10X (Part No 18-1092) obtained from Sartorius. 600nm polyglycerol-coated fluorescent nanodiamonds (FNDs) were purchased from Adámas Nanotechnologies. Triton X-100 lysis buffer, pH 7.4 (Cat # J62289, Alfa Aesar) purchased from Thermo Scientific. Polystreptavidin test line nitrocellulose half-strips were purchased from Global Access Diagnostics (UK). N,N-Dimethylformamide (Cat#227056), N,N-Disuccinimidyl carbonate (Cat#43720), IGEPAL CA-630 (Cat#I8896), Trizma hydrochloride (Cat#T3253), Trizma base (Cat#T6066), Casein Hammarstein Bovine (Cat#E0789) were purchased from Sigma Aldrich. NaCl (Cat#27810.295) and Tween20 (Cat#437082Q) purchased from VWR chemicals. SARS-CoV-2 nucleocapsid antibodies (40143-R001, 40143-R001-B, 40143-R004, 40143-R040, 40143-MM08), SARS-CoV-2 biotinylated recombinant nucleocapsid protein (40588-V08B-B), and MERS-CoV (40068-V08B), HCoV-OC43 (40643-V07E), HCoV-299E (40640-V07E), HCoV-NL63 (40641-V07E), HCoV-HKU1 (40642-V07E) recombinant nucleocapsid protein were purchased from Sino Biological. SARS-CoV-2 recombinant nucleocapsid protein was provided by Peter Cherepanova (Francis Crick Institute) and Prof Ciara O'Sullivan at Universitat Rivira i Virgili (URV). AbCR3009 and AbCR3018 were provided by Laura McCoy at UCL Division of Infection and Immunity. The following reagents were obtained through BEI Resources; NIAID, NIH: SARS-Related Coronavirus 2, Isolate USA-WA1/2020, Gamma-Irradiated, NR-52287, contributed by the Centers for Disease Control and Prevention, and NIH: SARS-Related Coronavirus 2, Isolate hCoV-19/USA/GA-EHC-2811C/2021 (Lineage B.1.1.529; Omicron

Variant), Gamma-Irradiated, NR-56496, contributed by Mehul Suthar. 40nm citrate gold nanoparticles (AuNPs) were purchased from Nanocomposix (SKU: AUCR40-5M). ACON Biotech FlowFlex SARS-CoV-2 Rapid Antigen Test (Self-Testing) 25 Tests (Ref L031-118Q5 PZN-17522027). Clinical Nasopharyngeal swab samples were obtained from UCLH. N1 2019-nCoV RUO kit (Integrated DNA Technologies Cat#10006713). SARS-CoV-2 synthetic RNA positive control (Twist Bioscience, control 51 cat#105346). In-house lateral flow test assembly: Nitrocellulose membrane CN95 (UniStart Sartorius part no. 1UN95ER050025WS), backing card 60mmx300mm purchased from Kenosha (KN-PS1060.45), sink pad: cellulose fiber 20cm x 30cm CFSP223000 (Millipore)

#### b) Running buffer formulation

A clean beaker was placed on a magnetic stir plate with a magnetic bar for continuous stirring. Formulation was prepared at 5X concentration of IGEPAL CA-630, Casein, and Tween20 in 50mM Tris-HCl + 100mM NaCl base buffer. 50mM TRIS-HCl solution was made up to pH 8 mixing appropriate amounts of Trizma Hydrochloride (MW: 157.60 g/mol) + Trizma Base (MW: 121.14 g/mol) (mg) in DI water, continuously stirring until fully dissolved. NaCl (Sigma) was added at 100mM in final solution volume, continuously stirring until fully dissolved. The 50mM Tris-HCL + 100mM NaCl buffer was split into two batches, one stored at 2-8°C until further use, and the second batch was used to prepare the 5X buffer. While continuously stirring, 10% w/v IGEPAL CA-630 was slowing added to the mixture, followed by 0.25% w/v Tween20. Next, 4% w/v Casein (Hammarstein bovine, Sigma Cat#E0789) was added, and the temperature was raised to 40°C monitored by a temperature probe, covered, and continuously stirred for 24hrs. Following, the buffer was aliquoted and frozen at -20°C. The 5X buffer was thawed and diluted 1:5 in base buffer (50mM Tris-HCL, pH8 +100mM NaCl) to make a 1X solution and stored at 4°C for lateral flow testing.

#### c) Lateral flow test line printing

CN95 nitrocellulose membrane at 2.5cm width (Sartorius) was laminated onto backing card (Kenosha) with 2.2cm absorbent pad (Millipore) and cut into 15mm card segments for printing (Biodot AD1520 dispenser). Antibodies were diluted from stock concentration to 1mg/mL in 1X PBS for printing at 1μL/cm, with test line printed 1cm from the bottom of the nitrocellulose membrane. Following printing, cards were placed in the oven to dry for 2hrs at

37°C. Strips were cut with strip cutter (ZQ2002, Shanghai Kinbio Tech. Co., Ltd.) at 3mm width. Strips were stored in a desiccated bag until use.

#### d) Gold nanoparticle (AuNP) functionalisation

The parameters for antibody physisorption were determined using 40 nm citrate AuNPs (Nanocomposix) at OD1 and swept across antibody concentrations from 8.7 to 70 μg/mL, in buffers ranging from pH 7.7 to 9.0. A salt stress test was applied and particle absorption spectra was measured using plate reader (SpectraMax i3, Molecular Devices) to check for aggregation and absorption properties. A 1mL aliquot of AuNPs was mixed with 200μL of AbMM08 at 35μg/mL in borate buffer pH 8. This mixture was incubated at room temperature for 1hr at 650RPM (Thermoshaker). Blocking was performed by adding 100μL of 1mg/mL BSA in $H_2O$ and left to shake for 30mins. The mixture was washed by centrifuge (3 washes at 10,000 rcf for 10mins) and resuspended in 5% BSA + 0.05% Tween20 in PBS.

### 2) Capture Antibody-Antigen Kinetic Analysis using the Forte Bio Octet

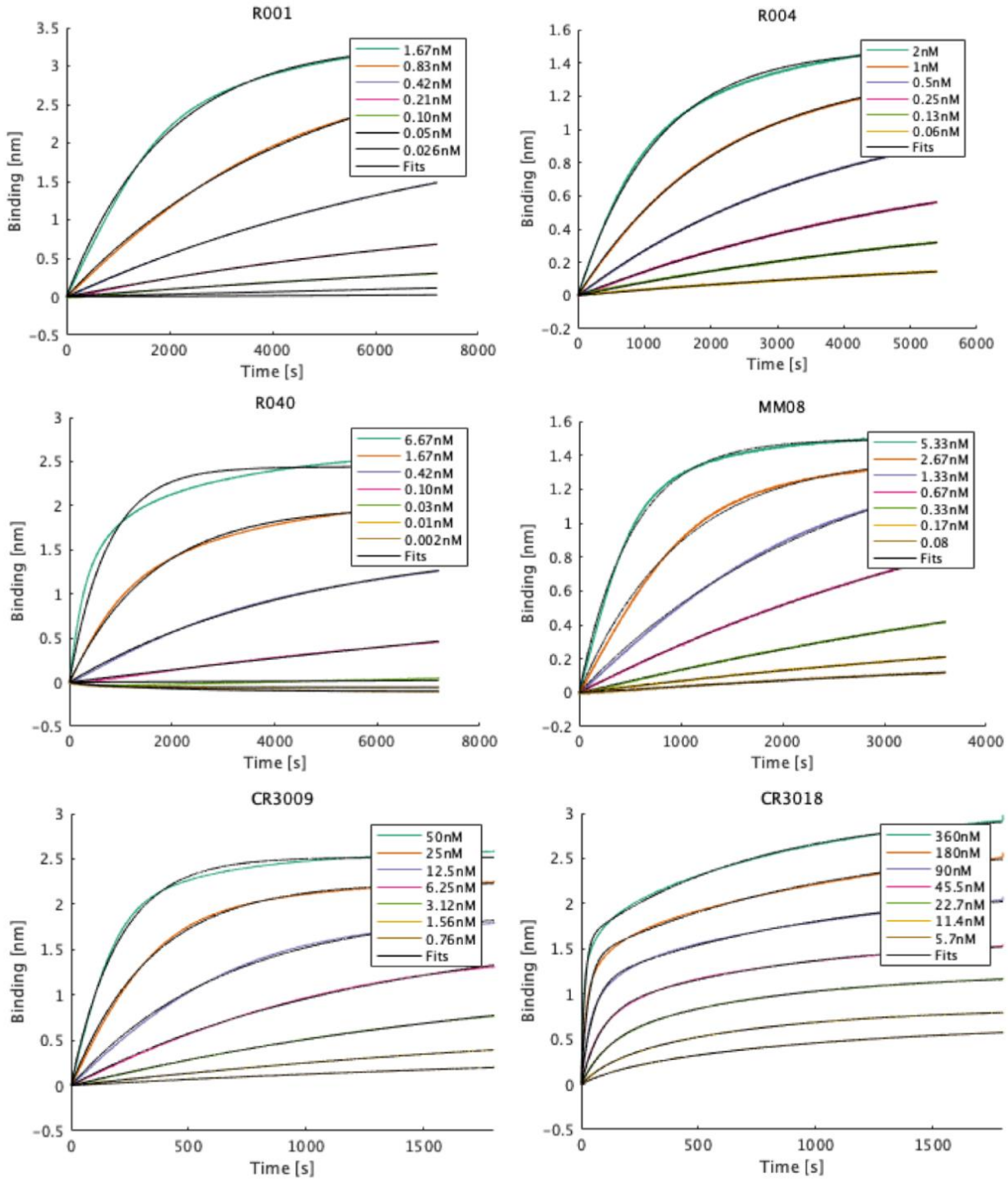

***Supplementary Information Fig. 1:*** *Association plot of intensity-time binding for different concentrations of target recombinant nucleocapsid protein for each antibody. Coloured lines represent test data and black lines represent fitted data from Langmuir fit. Antibody CR3018 showed bivalent binding characteristics. Binding curves provided by Sino Biological for antibodies R001 and R040 resulted in* $K_D$ *values of 0.02 nM and 0.01 nM, respectively*[1,2].

| Capture Antibody (Manufacturer, ID) | $K_D$ (nM) [95% CI] | $K_{on}$ (sM) [95% CI] | $K_{off}$ ($s^{-1}$) [95% CI] |
|---|---|---|---|
| **R001** (Sino Biological, RRID: AB_2827974) | 0.34 [0.19-0.55] | $3.0 \times 10^{5}$ [2.6-3.5] | $2.0 \times 10^{-6}$ [-33-37] |
| **R004** (Sino Biological, RRID: AB_2827975) | 0.22 [0.17-0.27] | $3.7 \times 10^{5}$ [3.5-4.0] | $8.5 \times 10^{-5}$ [6.0-11] |
| **R040** (Sino Biological, RRID: AB_2827976) | 0.25 [0.12-0.49] | $1.7 \times 10^{5}$ [-1.79-5.16] | $2.3 \times 10^{-4}$ [-12-16] |
| **MM08** (Sino Biological, RRID: AB_2827978) | 0.19 [0.091-0.36] | $3.6 \times 10^{5}$ [3.2-3.9] | $1.3 \times 10^{-5}$ [-6.7-9.4] |
| **CR3009** (In-house Dr Laura E McCoy, clone ID 03-009[3]) | 3.3 [2.7-3.9] | $9.7 \times 10^{4}$ [8.8-11] | $2.6 \times 10^{-4}$ [0.71-4.4] |
| **CR3018** (In-house Dr Laura E McCoy, clone ID 03-018[3]) | 69 [62-76] | $1.6 \times 10^{5}$ [1.6-1.7] | $5.8 \times 10^{-3}$ [5.0-6.7] |

***Supplementary Information Table 1.*** *Summary of capture antibody-antigen kinetic parameters including equilibrium dissociation constant, $K_D$, association rate, $k_{on}$, and dissociation rate, $k_{off}$ obtained using BLI*

3) Performance evaluation with recombinant antigen

a) Comparison of two different sources of recombinant nucleocapsid protein

Performance of the two different antigen sources (Sino biological and O'Sullivan Lab, URV) were compared at select test concentrations with the FND SARS-CoV-2 antigen assay during initial assay development.

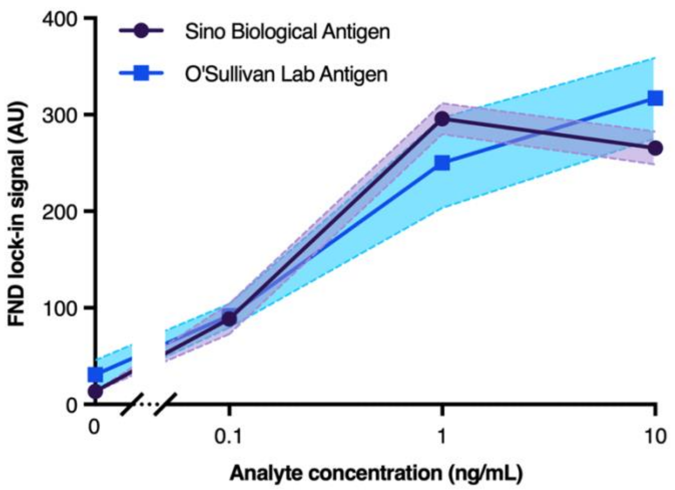

*Supplementary Information Fig. 2: Comparison of two different sources of nucleocapsid protein. Performance with commercially available nucleocapsid protein (Sino Biological Inc.), and nucleocapsid protein developed in-house by O'Sullivan Lab, tested on the FND SARS-CoV-2 antigen assay showing similar performance across the same antibody pair (t-test paired p-value 0.7621, T=0.3315, DF=3).*

b) Statistical comparison of assay LoD fitting models

| | 4PL | Exponential | Langmuir | Stretch exponential |
|---|---|---|---|---|
| FND Recombinant nucleocapsid protein LoD | | | | |
| RMSE | 0.066 | 0.056 | 0.056 | 0.057 |
| AICc | -120.1 | -137.2 | -137.0 | -134.8 |
| AuNP Recombinant nucleocapsid protein LoD | | | | |
| RMSE | 0.048 | 0.046 | 0.046 | 0.043 |
| AICc | -121.3 | -125.7 | -125.1 | -129.8 |

*Supplementary Information Table 2: A statistical method for determining the limit of detection (LoD) was employed based on methods reported in Holstein et al.,[4] and adapted by Miller et al.[5] Comparison of suitability of LoD fitting models considered[6]: the four-parameter logistic regression, exponential, Langmuir adsorption model, and stretch exponential. A comparison of root mean squared errors (RMSE) and small-sample corrected Akaike Information Criterion (AICc) is shown for all 4 models. All models show excellent RMSE. The 4PL model was the least sensitive. The AICc values show the exponential fit was the best model.*

### c) Comparison of assay sensitivities reported in literature

| Reference | Sensor type | LoD | Clinical evaluation high viral load (Ct$\leq$**25 or $10^6$ copies/mL)** sensitivity |
|---|---|---|---|
| Chen et al. (2022)[7] | Multiplex fluorescent quantum dot LFT | 10 pg/mL | No clinical samples tested |
| Grant et al. (2021)[8] Bachman et al. 2021)[9] | Latex bead LFT | 200 $TCID_{50}$/mL | 92% (n=72) |
| Wang et al. (2021)[10] | Magnetic quantum dot LFT | 1 pg/mL (direct mode) | No clinical samples tested |
| Wei-Wen Hsiao et al. (2022)[11] | Fluorescent nanodiamond LFT (magnetic modulation) | 1.94 ng/mL | No clinical samples tested |
| Gupta et al. (2023)[12] | Fluorescent gold nanorod LFT | 212 pg/mL | 97.5% (n=40) |
| This work | Fluorescent nanodiamond LFT (MW modulation) | 0.67 pg/mL | 100% (n=30) |

***Supplementary Information Table 3****: Summary of SARS-CoV-2 Ag-LFTs targeting nucleocapsid protein reported in literature with their assay limit of detection.*

4) Extended clinical sample analysis

a) Clinical sensitivity and specificity ROC summary

| Area under the ROC curve | All samples | Samples Ct ≤ 30 | Samples Ct ≤ 25 |
|---|---|---|---|
| Area | 0.92 | 0.98 | 1.00 |
| Std. Error | 0.03 | 0.02 | 0.00 |
| 95% confidence interval | 0.86 to 0.99 | 0.94 to 1.00 | 1.000 to 1.000 |
| P value | <0.0001 | <0.0001 | <0.0001 |
| Data | | | |
| Controls (Negative) | 37 | 37 | 37 |
| Patients (Signal) | 53 | 41 | 30 |

***Supplementary Information Table 4: Results summary of Area under the ROC curve***. *ROC curve analysed for all samples (n=53) and sub-categories for Ct ≤ 30 and Ct ≤ 25.*

b) Clinical sample matrix stratification by nasal and nasopharyngeal swab type

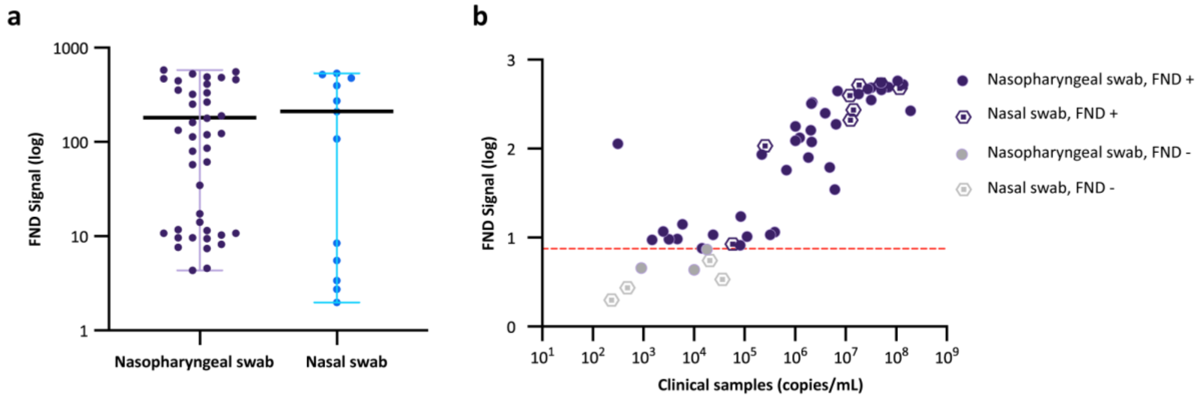

***Supplementary Information Fig. 3: (a)*** *Box plot of positive clinical sample FND results stratified by nasopharyngeal swab and nasal swab type showing mean FND signal (black line). No significant difference between mean FND signal was observed (t-test p-value=0.6457, T=0.4625, DF=51).* ***(b)*** *Visual representation of FND results by clinical sample viral load and swab type showing distribution of viral loads in nasal swabs and nasopharyngeal swabs (no significant difference between mean viral load across sample type determined by t-test, p-value=0.9886, T=0.01432, DF=51).*

### 5) Estimating AuNP Ag-LFT clinical sensitivity compared to FND Ag-LFT sensitivity evaluation

In order to calculate the clinical sensitivity of the AuNP assay, a relative threshold compared to FNDs was calculated. This threshold was taken as the FND signal for a sample with a recombinant antigen concentration corresponding to the fitted LoD of the AuNP assay. The signal y-value ($\log_{10}$) was calculated based on the following equation:

$$y = log_{10}\left(a + b * \left(1 - exp\left(-\frac{10^{[LOD_{AuNP}]} - 2}{c}\right)\right)\right)$$

where $y$ is the FND signal corresponding to the AuNP LoD, $LOD_{AuNP}$ is the AuNP LoD, and $a$, $b$, and $c$ are parameters fitted in Fig. 2a. The threshold was applied to clinical sample data (Fig. 3b) to estimate clinical sensitivity of the AuNP assay.

| | High viral load ($\geq 10^6$ copies/mL) | moderate viral load ($10^5$-$10^4$ copies/mL) | Full range ($10^2$-$10^8$ copies/mL) |
|---|---|---|---|
| FND sensitivity | 100% | 90% | 87% |
| AuNP sensitivity | 93% | 20% | 57% |

***Supplementary Information Table 5****: Summary of assay sensitivity grouped by viral load of clinical samples using extrapolated threshold calculated for the AuNP assay. High viral load samples n=29, moderate viral load samples n=10, full range n=53. Sensitivities were calculated non-cumulatively, as opposed to Fig. 3a grouped by Ct with cumulative sensitivities. The sensitivities calculated within each viral load category were used for modelling analysis detailed in Supplementary Information Section 6 and Fig.4c.*

### 6) Modelling clinical assay performance from infection dynamics of SARS-CoV-2 human challenge infection

Infection dynamics data from the SARS-CoV-2 human challenge trial[13] were used to calculate the expected clinical performance of the FND and AuNP assay over the course of infection to estimate the time from infection to detection for each assay. The challenge infection data provides the true days of infection, independent of symptom onset. The data plot is extracted from Killingley et al.[13] Fig. 2a, which represents mean viral load and standard error of the mean in nasal swab samples taken twice daily from the participants (n=18). The study found a median of ~2.24 days [95% CI: 1.67-3.17] to initial quantifiable detected virus with RT-qPCR at a threshold set at $10^3$ copies/mL. We calculate the mean day of initial detection from patient samples RT-PCR at ~2.08 days.

First, we used a symmetric Bayesian linear regression with errors-in-variables (x and y) to fit FND signal ($\log_{10}$) and viral load ($\log_{10}$(copies/mL))[14]. This used Gaussian prior distributions for the gradient and intercept, both with a mean of 0 and a standard deviation of 10. The log likelihood function uses the orthogonal residuals from the model, weighted by the variance in the orthogonal direction:

$$\text{log likelihood} = \sum_{i=1}^{n}\left[-\frac{1}{2}\ln(2\pi\sigma_i^2) - \frac{d_i^2}{2\sigma_i^2}\right]$$

where $d_i$, is the orthogonal residual, and $\sigma_i^2$ is the combined variance from uncertainty in x and y:

$$d_i = \frac{y_i - \hat{y}_i}{\sqrt{1 + b^2}}$$

where $y_i$ is a measured $y$ value, $\hat{y}_i$ is the $y$ value for a given $x_i$ predicted by the model: $\hat{y}_i = a + bx_i$ with y-intercept $a$, and gradient $b$.

$$\sigma_i^2 = \frac{\sigma_{y_i}^2 + b^2\sigma_{x_i}^2}{\sqrt{1 + b^2}}$$

where $\sigma_{y_i}^2$ is the $y$-variance and $\sigma_{x_i}^2$ is the $x$-variance of a point $(x_i, y_i)$.

This model was then used to calculate the equivalent copies/mL values and associated uncertainty for the FND signal LoD threshold, and relative FND signal for the AuNP LoD (see Supplementary Information 4). The uncertainty was calculated by propagation of errors:

$$x_{est} = \frac{y_t - a}{b}$$

$$\sigma_{x_{est}} = \sqrt{\left(\frac{\sigma_{y_t}}{b}\right)^2 + \left(\frac{\sigma_a}{b}\right)^2 + \left(\frac{(y_t - a)\sigma_b}{b^2}\right)^2}$$

where $x_{est}$ is the estimated copies/mL for a FND signal threshold $y_t$.

The Metropolis-Hastings algorithm is used to sample from the posterior distribution[15].

The FND threshold was 5.6 $\times 10^4$ copies/mL [95% credible interval: 4.2 $\times 10^4$ to 7.5 $\times 10^4$] and AuNP threshold was 3.5 $\times 10^6$ copies/mL [95% credible interval: 2.3 $\times 10^6$ to 5.1 $\times 10^6$].

These thresholds were then plotted on the infection dynamics graph (Killingley et al., Fig. 2a) and interpolated to find the estimated timepoint (days) to initial detectable virus across each assay (Fig. 4a). The day of initial detection for the patient with the mean viral load was calculated for both the FND and AuNP assay with 95% confidence intervals, accounting for error in the viral load thresholds. Following, the difference between the mean days with standard error was used to indicate the average window for earlier detection with FNDs. We then used the distribution of patient viral loads over time (Killingley et al.) along with the uncertainty in viral load thresholds to calculate the percentage of detectable patients over time for each assay and, correspondingly, the FND diagnostic advantage in days against patient detection percentage.

### 7) Calculating expected clinical assay performance by day of infection in a clinical symptomatic cohort

This analysis uses calculated sensitivity of FND and AuNP assays across a range of viral loads to show the approximate number of infected patients that could be detected per day. This data is adapted from Frediani et al.,[16] that presented a large clinical study of viral loads in the days following symptom onset. The proportion of patients with high viral load samples ($Ct \leq 25$), moderate viral load ($25 < Ct \leq 30$), and low viral load ($Ct > 30$) sampled on each day was used to estimate the patients detected by the FND and AuNP assay per day. The calculated assay sensitivities by viral load, shown in Supplementary Information Table 5, were applied proportionately to the number of patients samples within the viral load range per day. This was used to determine the percentage of patients that would be detected with the FND and AuNP assay on each day of infection (Fig. 4c).

### 8) Estimating number of patients detected in a single day at peak pandemic

To put the spin-enhanced LFT diagnostic advantage in context of patient numbers at the population level, we applied the respective assay sensitivity to the total number of confirmed COVID-19 cases in a single day at peak pandemic to estimate how many more patients the FND assay may have detected than the AuNP assay. In this case, the daily reported case number does not include stratification of individual patient data by day of symptom onset or viral load, therefore we applied the total assay sensitivity for this calculation. The sensitivity was 87% and 57% for the FND assay and AuNP assay, respectively (Supplementary Information Table 5). This analysis assumes that the sensitivity calculated from our clinical sample data set is generally representative of the patient population.

At the height of the Omicron wave in the UK on ~5 Jan 2022, the daily confirmed cases reached ~229,901 (Supplementary Information Fig. 4) (https://ourworldindata.org/covid-cases), in which approximately 69,400 more patients may have been detected by the FND assay than the AuNP assay. This assumes our clinical sample set is representative of the population viral load distribution at the peak of the Omicron wave.

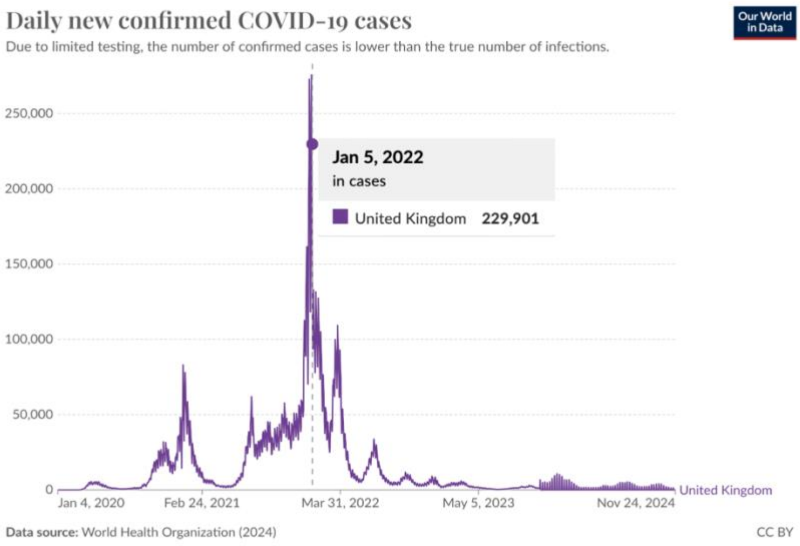

***Supplementary Information Fig. 4:*** *WHO data tracking daily new confirmed COVID-19 cases in the United Kingdom over the course of the pandemic (Jan 2020 to Nov 2024). Marker indicated on Jan 5$^{th}$ corresponds to the Omicron peak reaching nearly 230,000 new cases in a single day. Graph downloaded from https://ourworldindata.org/covid-cases.*

### 9) Assessing performance retention transitioning to an integrated point-of-care platform

The assay presented has been designed for greater flexibility in design and optimisation throughout the initial stages of development in a lab-based setting. This section addresses the feasibility of retaining similar performance in a fully integrated lateral flow platform. These considerations include (i) adequate reaction kinetics for on-pad reagent delivery in cassette format (ii) immediate read-out of strip following 15min run time (without drying) for fast time-to-results and (iii) feasibility of cost-effective, portable fluorescence read-out device.

#### a) Reaction kinetics study: Demonstrating equivalent assay performance with direct strip application.

This study shows feasibility of eliminating the 10-minute incubation of reagents prior to adding the lateral flow strip, which more closely represents the binding interactions that occur in a cassette-based lateral flow format as complexes are formed and captured immediately during flow up the membrane. Strips were tested with a wait time from 0-10 mins at 1 min intervals, evaluating the impact on the SNR from a low positive (31 pg/mL) and negative sample (n=3) over time. A low analyte concentration (31 pg/mL) was chosen for this study to reflect a case where complex formation/binding is less favourable and therefore more likely to observe potential impact on performance than at high concentrations where binding time has diminished impact.

The assay was performed as follows: 5μL of the FNDs (26 fM) + 49μL of sample diluted in running buffer + 1μL of bAb01 (513nM) was added to wells in a 96-well plate and allowed to bind for 0-10 mins. Strip were added to the wells and run for 15 mins, followed by test line read-out of dry strips. Time 0 mins corresponds to application of the lateral flow strip immediately after adding reagents to the wells and 10 min wait time is representative of the protocol used throughout this work. No significant difference in SNR was observed across the time intervals tested (Supplementary Information Fig. 5), indicating the complex formation in solution does not impact effective capture at the test line. In future, eliminating the "binding time" step for this assay can reduce assay time to the standard ~15 min lateral flow test run time without impacting performance.

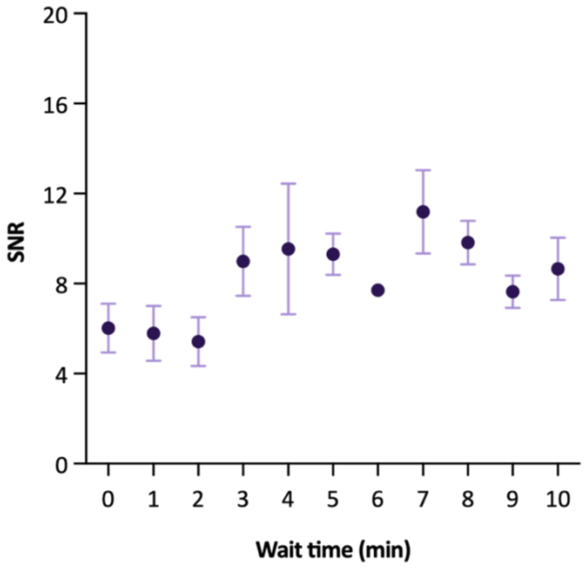

**Supplementary Information Fig. 5: (a)** The SNR of negative (n=3) and low positive (31 pg/mL, n=3) tested at 1 min time intervals where at time 0, the lateral flow strips were ran immediately after addition of capture/detection reagents. Data points represent mean SNR with error bars representing the standard error from the mean, showing no significant difference between mean SNRs across the time interval (one-way ANOVA, Tukey's multiple comparisons, p-value=0.1479, F= 1.68, DF=32) and a gradient not significantly different from zero using a linear regression fit (95%CI: -0.0089 to 0.67).

#### b) Impact of lateral flow test strip drying on FND lock-in signal

This short study looked at the effect of reading wet lateral flow strips on the resonator-based read-out platform. This would allow for read-out of lateral flow strips immediately following running, reducing the total assay time to fit the point-of-care criteria. The key potential confound for wet strips is the shift in the resonator's resonant frequency (water has a high dielectric constant).

Test strips from a positive (n=2) and negative (n=3) sample were measured using lock-in analysis at 1-minute time intervals from 0-60 mins from the end of strip running. We observed a ~5 point reduction in lock-in values in the first ~10 minutes of read-out, likely due to FNDs still washing up the strip. This happens in all LFTs, not just nanodiamond LFTs. This is a higher proportion of the final lock-in value for the negatives, as the signal is lower, so there is a net effect on SNR: lower immediately after reading. It will have a minimal effect on sensitivity and can be mitigated with a differential readout (normalising to strip background).

Over the subsequent period of about 5-minutes, the signals rise in both positive and negatives tests as the strips dry. The overall effect on SNR is therefore zero. This can therefore be mitigated by taking ratiometric measurements of test line and background (analogous to test line and signal), or test line and control line. Finally, the resonator could be tuned to account for the changes in resonant frequency with wet strips (either in real-time or preset).

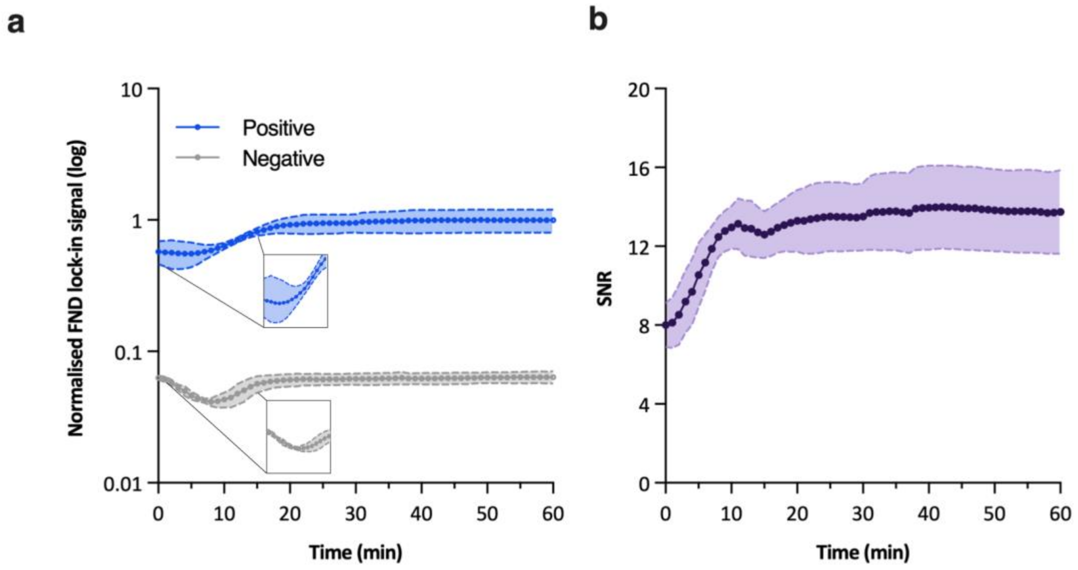

***Supplementary Information Fig. 6: (a)*** *The normalised FND lock-in signal of n=3 negatives (grey) and n=2 positives (blue) measured at 1-minute intervals over 60 minutes, shaded region showing standard deviation. The effect of wetting reduced lock-in signal of positive samples by ~40% of its maximum value at time 0, with a coefficient of variation (CV) = 18.3% across all time points. Minimal effect on the negatives was observed CV=11.2% across all time points (no error bars shown for error <0.1). Inset shows region from 0-15 minutes on a linear scale, where an initial decrease in lock-in signal is observed, likely due to FNDs washing away while the strip is still wet and solution is still moving across the strip. This is then counterbalanced by the strip drying, increasing signal again. In the positive sample, many of these particles bind to the test line rather than flowing past, giving a smaller dip.* ***(b)*** *The signal-to-noise ratio was calculated from the means in (a), with a minimum SNR of 8 (±1.1) at time 0 and reaching a plateau ~11 mins with SNR=13 (±1.2). The SNR has variation of 11.2% across 60 mins.*

### c) Estimated costs of portable fluorescent reader

The feasibility of a portable fluorescent reader with modulation capabilities is a key component to the translation of the FND platform to a point-of-care setting. A portable, smartphone-based read-out system meets point-of-care criteria using cost effective 3D-printed housing, miniaturised electronics, and small resonator components allow for integration into a portable device that can be deployed in the field and at the point-of-care including doctors' offices and pharmacies. A costing summary of a prototype FND device is shown in Supplementary Information Table 6.

| Device | Description | Cost (£) |
|---|---|---|
| FND prototype smartphone reader | Resonator | 6 |
| | 30mW laser | 93 |
| | Lens & filters | 221 |
| | Cables & 3D printed material | 25 |
| | Voltage controlled oscillator & chip | 65 |
| | amplifier | 137 |
| | Power bank | 20 |
| | Smartphone (LG-G7) | 345 |
| | **Total** | **912** |

***Supplementary Information Table 6:*** *Estimated costing of FND portable reader with smartphone connectivity in development.*

### 10) Supplementary Information References

1. Sino Biological. *Datasheet Catalog Number 40143-R001*. https://www.sinobiological.com/antibodies/cov-nucleocapsid-40143-r040 (2020).
2. Sino Biological. *Datasheet Catalog Number 40143-R040*. https://www.sinobiological.com/antibodies/cov-nucleocapsid-40143-r040 (2020).
3. van den Brink, E. N. *et al.* Molecular and biological characterization of human monoclonal antibodies binding to the spike and nucleocapsid proteins of severe acute respiratory syndrome coronavirus. *J Virol* **79**, 1635–1644 (2005).
4. Holstein, C. A., Griffin, M., Hong, J. & Sampson, P. D. Statistical Method for Determining and Comparing Limits of Detection of Bioassays. *Anal Chem* **87**, 9795–9801 (2015).
5. Miller, B. S. *et al.* Spin-enhanced nanodiamond biosensing for ultrasensitive diagnostics. *Nature* **587**, 588–593 (2020).
6. Miller, B. S. *et al.* Sub-picomolar lateral flow antigen detection with two-wavelength imaging of composite nanoparticles. *Biosens Bioelectron* **207**, 956–5663 (2022).
7. Chen, W. *et al.* An integrated fluorescent lateral flow assay for multiplex point-of-care detection of four respiratory viruses. *Anal Biochem* **659**, 114948 (2022).
8. Grant, B. D. *et al.* A SARS-CoV-2 coronavirus nucleocapsid protein antigen-detecting lateral flow assay. *PLoS One* **16**, e0258819 (2021).
9. Bachman, C. M. *et al.* Clinical validation of an open-access SARS-COV-2 antigen detection lateral flow assay, compared to commercially available assays. *PLoS One* **16**, e0256352 (2021).
10. Wang, C. *et al.* Ultrasensitive and Simultaneous Detection of Two Specific SARS-CoV-2 Antigens in Human Specimens Using Direct/Enrichment Dual-Mode Fluorescence Lateral Flow Immunoassay. *ACS Appl Mater Interfaces* **13**, 40342–40353 (2021).
11. Wei-Wen Hsiao, W. *et al.* Fluorescent nanodiamond-based spin-enhanced lateral flow immunoassay for detection of SARS-CoV-2 nucleocapsid protein and spike protein from different variants. *Anal Chim Acta* **1230**, 340389 (2022).
12. Gupta, R. *et al.* Ultrasensitive lateral-flow assays via plasmonically active antibody-conjugated fluorescent nanoparticles. *Nat Biomed Eng* **7**, 1556–1570 (2023).
13. Killingley, B. *et al.* Safety, tolerability and viral kinetics during SARS-CoV-2 human challenge in young adults. *Nat Med* **28**, 1031–1041 (2022).
14. Dellaportas, P. & Stephens, D. A. Bayesian Analysis of Errors-in-Variables Regression Models. *Biometrics* **51**, 1085–1095 (1995).
15. Hastings, W. K. Monte Carlo sampling methods using Markov chains and their applications. *Biometrika* **57**, 97–109 (1970).
16. Frediani, J. K. *et al.* The New Normal: Delayed Peak SARS-CoV-2 Viral Loads Relative to Symptom Onset and Implications for COVID-19 Testing Programs. *Clinical Infectious Diseases* **78**, 301–307 (2024).
